# International prevalence patterns of low eGFR in adults aged 18-60 without traditional risk factors from population-based cross-sectional studies: a disadvantaged populations eGFR epidemiology (DEGREE) study

**DOI:** 10.1101/2024.06.24.24309380

**Authors:** Charlotte E Rutter, Mary Njoroge, Phil Cooper, Prabhakaran Dorairaj, Vivekanand Jha, Prabhdeep Kaur, Sailesh Mohan, Ravi Raju Tatapudi, Annibale Biggeri, Peter Rohloff, Michelle H Hathaway, Amelia Crampin, Meghnath Dhimal, Anil Poudyal, Antonio Bernabe-Ortiz, Cristina O’Callaghan-Gordo, Pubudu Chulasiri, Nalika Gunawardena, Thilanga Ruwanpathirana, SC Wickramasinghe, Sameera Senanayake, Chagriya Kitiyakara, Marvin Gonzalez-Quiroz, Sandra Cortés, Kristina Jakobsson, Ricardo Correa-Rotter, Jason Glaser, Ajay Singh, Sophie Hamilton, Devaki Nair, Aurora Aragón, Dorothea Nitsch, Steven Robertson, Ben Caplin, Neil Pearce, the DEGREE Study Group

## Abstract

The <u>d</u>isadvantaged populations <u>eG</u>F<u>R</u> (estimated glomerular filtration rate) <u>e</u>pid<u>e</u>miology (DEGREE) study was designed to gain insight into the burden of chronic kidney disease (CKD) of undetermined cause (CKDu) using standard protocols to estimate the general-population prevalence of low eGFR internationally.

We estimated the age-standardised prevalence of eGFR<60ml/min/1.73m^2^ in adults aged 18-60, excluding participants with commonly known causes of CKD, i.e., ACR>300mg/g or equivalent, or self-reported or measured hypertension or diabetes (eGFR<60_[absent HT,DM,high ACR]_), and stratified by sex and location. We included population-representative surveys conducted around the world that were either designed to estimate CKDu burden or were re-analyses of large surveys.

There were 60 964 participants from 43 areas across 14 countries, with data collected during 2007-2023. The highest prevalence was seen in rural men in Uddanam, India (14%) and Northwest Nicaragua (14%). Prevalence above 5% was generally only observed in rural men, with exceptions for rural women in Ecuador (6%) and parts of Uddanam (6-8%), and for urban men in Leon, Nicaragua (7%). Outside of Central America and South Asia, prevalence was below 2%.

These observations represent the first attempts to estimate the prevalence of eGFR<60_[absent HT,DM,high ACR]_ around the world, as an estimate of CKDu burden, and provide a starting point for global monitoring. It is not yet clear what drives the differences, but available evidence to date supports a high general-population burden of CKDu in multiple areas within Central America and South Asia, although the possibility that unidentified clusters of disease may exist elsewhere cannot be excluded.

**Lay summary:** In recent decades there have been reports of epidemics of chronic kidney disease (CKD) killing young men in Central America and South Asia. These cases do not involve the commonly known causes of CKD such as diabetes, so they are known as CKD of unknown cause (CKDu). To understand the size and extent of the problem around the world, we included data from studies that measured kidney function from 43 areas across 14 countries (60 964 people). We calculated the prevalence of poor kidney function in working-age men and women in urban and rural areas, in those without indicators of the commonly known causes of CKD. The most affected groups were rural men in Uddanam, India (14%), and Northwest Nicaragua (14%). Low prevalence (<2%) was seen in included areas outside of Central America and South Asia. These findings are important to direct future research, give clues to the possible causes of CKDu, and as a starting point for global monitoring.

## Introduction

Globally, chronic kidney disease (CKD) is most commonly associated with diabetes, hypertension, other cardiovascular diseases, glomerulonephritis, genetic or congenital abnormalities, or urological diseases. However, there is an increasing recognition of forms of progressive CKD which are not associated with these known risk factors, and which are mostly affecting the working-age populations in low- and middle-income countries (LMICs).^1,2^ This clinical syndrome has been termed CKD of undetermined cause (CKDu); other names used include CKD of non-traditional cause (CKDnt), Mesoamerican Nephropathy (MeN), Uddanam nephropathy, and chronic interstitial nephritis of agricultural communities (CINAC). Over the last few decades, clusters of CKDu have been reported in Central America,^3^ Mexico,^4^ India,^5^ and Sri Lanka.^6^ Other reports have suggested that similar patterns may be occurring in other regions of the world, but it is only recently that efforts have increased to undertake comparable population surveys in working-age populations elsewhere in LMICs.

Perhaps the most clearly established risk factor/epidemiological association in both Central America and South Asia, is that CKDu is more common among men engaged in manual labour in hot climates, particularly in agricultural communities.^7^ In Central America, CKDu occurs frequently in sugar cane workers but also in other occupational groups, including other agricultural workers, fishermen, miners, brick kiln and construction workers;^8^ it also occurs, albeit at a lower frequency, in women, most of whom have not reported working in agriculture. In common with historical endemic kidney diseases such as Balkan Nephropathy,^9^ the absence of substantial albuminuria or haematuria, alongside geographical clustering, supports a primarily tubular-interstitial disease, and potentially a causal role for environmental exposure(s). Many specific potential causes related to agriculture have been suggested for CKDu. Heat/dehydration, pesticides, and heavy metals are the main hypotheses proposed for Central America, whereas in South Asia the emphasis has been on the possible roles of water contamination by metals and/or pesticides.^10–12^

In the past, international comparisons have played a key role in identifying possible causes of chronic disease.^13^ For example, many of the discoveries on the causes of cancer (e.g., human papilloma virus and cervical cancer) have their origins, directly or indirectly, in the systematic international comparisons of cancer incidence conducted in the 1950s and 1960s. Hypotheses generated from these studies were investigated in more depth in further studies.^14^ A more recent example is the International Study of Asthma and Allergies in Childhood (ISAAC), a standardised protocol to estimate the prevalence of asthma internationally,^15,16^ which has now evolved into the Global Asthma Network.^17,18^ This has led to a greater understanding of the possible causes of asthma globally, as well as the creation of a large international network of researchers.

We have proposed a similar approach involving a simple and practical protocol to describe distributions of kidney function, using the estimated glomerular filtration rate (eGFR), in disadvantaged communities globally: the <u>d</u>isadvantaged populations <u>eGFR e</u>pid<u>e</u>miology (DEGREE) study. The DEGREE protocol was explicitly developed for general population-based surveys.^19^ It was noted that the same methodology could be used in other contexts (e.g., workforce surveys), but the current paper focusses on population surveys.

As the causes of CKDu are unknown, diagnosis is often made by exclusion of known causes of kidney disease. The DEGREE protocol uses pragmatic criteria (absence of diabetes, hypertension, or heavy proteinuria) to estimate the prevalence of low eGFR unrelated to known causes of kidney disease (with the latter features being common in most forms of glomerular diseases). This enables standardised comparisons across multiple centres and is intended to identify population patterns, rather than diagnose CKDu in individuals.

We here report the first findings from the DEGREE study, involving 60 964 participants with complete data from 19 studies across 43 areas in 14 countries, with date of data collection varying by study between 2007 and 2023 (Table 1). These are primarily in LMICs, plus one study in rural Italy, another in Chile, and publicly available data from England and the United States as reference points for comparison.

**Table 1:** Characteristics of study areas and samples.

| Country | Study type | Rationale | Area name | Urban %<sup>a</sup> | Climate | Survey dates | Survey season | Source Population | Overall response rate % | Sample information<sup>b</sup> |  |  |
| --- | --- | --- | --- | --- | --- | --- | --- | --- | --- | --- | --- | --- |
|  |  |  |  |  |  |  |  |  |  | n | male % | age, years median (IQR) |
| **Chile** | Reuse of population survey<sup>37</sup> | Proposed risk factors | Molina | 69 | Mediterranean | Oct22-Nov23 | All year | 45 976 | 92 | 476 | 41 | 53 (48, 57) |
| **Ecuador** | CKD focused study<sup>38</sup> | DEGREE registered | Miguelillo, Manabi Province | 0 | Tropical | Jul21-Sep21 | Dry | 14 164 | 61 | 754 | 41 | 39 (28, 49) |
| **England** | HSE 2016 – Reuse of population survey<sup>21</sup> | Reference | England | 83 | Temperate | 2016 | All year | 56 000 000 | 59 | 2 135 | 42 | 44 (34, 52) |
| **Guatemala** | CKD focused study<sup>39</sup> | DEGREE registered | Tecpán, Chimaltenango | 0 | Temperate Highland Tropical | Jun18-Oct19 | All year | 85 000 | 58 | 336 | 34 | 34 (24, 47) |
|  |  |  | San Antonio Suchitepéquez | 0 | Tropical Wet | Jun18-Oct19 | All year | 52 000 | 69 | 318 | 34 | 33 (25, 45) |
| **India** | 1. CARRS - Reuse of population survey<sup>40</sup> | DEGREE registered | Chennai | 100 | Tropical | Oct10-Nov11 | All year | 4 680 000 | 92 | 5 366 | 43 | 39 (31, 47) |
|  |  |  | Delhi | 100 | Semi-arid | Oct10-Nov11 | All year | 16 300 000 | 96 | 3 564 | 49 | 42 (35, 50) |
|  | 2. ICMR-CHD – Reuse of population survey<sup>41</sup> | DEGREE registered | Delhi | 100 | Semi-arid | Aug11-Jan12 | Rainy, autumn, winter | 16 300 000 | Not reported | 1 888 | 44 | 41 (35, 48) |
|  |  |  | Faridabad | 0 | Semi-arid | Aug11-Jan12 | Rainy, autumn, winter | 90 000 | Not reported | 1 413 | 45 | 42 (36, 49) |
|  | 3. UDAY - Reuse of population survey<sup>42</sup> | DEGREE registered | Sonipat | 50 | Semi-arid | Jul14-Dec14 | Rainy, autumn, winter | 203 000 | 90<sup>c</sup> | 4 126 | 44 | 44 (37, 51) |
|  |  |  | Vizag | 50 | Tropical | Jul14-Dec14 | Rainy, autumn, winter | 275 000 |  | 4 209 | 44 | 43 (35, 50) |
|  | 4. Uddanam - CKD focused study<sup>43</sup> | Reported high CKDu area | Kanchili | 0 | Hot Tropical | Jun18- Dec19 | Summer, winter | 66 657 | 85<sup>c</sup> | 317 | 47 | 43 (35, 51) |
|  |  |  | Kaviti | 0 | Hot Tropical | Jun18-Dec19 | Summer, winter | 75 974 |  | 212 | 48 | 43 (35, 51) |
|  |  |  | Mandasa | 0 | Hot Tropical | Jun18-Dec19 | Summer, winter | 82 699 |  | 200 | 48 | 42 (33, 50) |
|  |  |  | Palasa | 0 | Hot Tropical | Jun18-Dec19 | Summer, winter | 97 551 |  | 362 | 51 | 44 (36, 51) |
|  |  |  | Sompeta | 0 | Hot Tropical | Jun18-Dec19 | Summer, winter | 78 908 |  | 443 | 46 | 42 (33, 51) |
|  |  |  | V_kothuru | 0 | Hot Tropical | Jun18-Dec19 | Summer, winter | 73 212 |  | 531 | 47 | 44 (35, 52) |
|  | 5. Prakasam – CKD focused study<sup>d</sup> | DEGREE registered | Kanigiri | 0 | Hot Tropical | Dec21-Feb22 | Winter | 1 780 | 84 | 1 052 | 40 | 39 (30, 49) |
| **Italy** | CKD focused study<sup>44</sup> | Reported high CKD area | Barga | 0 | Temperate | Jun21-Mar22 | Summer, autumn, winter | 9 574 | 92<sup>e</sup> (or 50<sup>f</sup>) | 301 | 43 | 47 (33, 54) |
| **Kenya** | CKD focused study<sup>45</sup> | DEGREE registered | Muhoroni East | 100 | Sub-tropical | Jul20-Nov20 | Dry | 3 740 | 85 | 260 | 53 | 34 (26, 43) |
|  |  |  | Owaga | 0 | Sub-tropical | Jul20-Nov20 | Dry | 3 769 | 87 | 242 | 47 | 36 (26, 46) |
|  |  |  | Tonde | 0 | Sub-tropical | Jul20-Nov20 | Dry | 3 045 | 98 | 233 | 49 | 36 (28, 45) |
| **Malawi** | CKD focused study<sup>46</sup> | DEGREE registered | Southern Karonga District | 0 | Sub-tropical | Jan18-Aug18 | Dry, rainy | 40 000 | 66 | 646 | 42 | 33 (24, 41) |
|  |  |  | Lilongwe | 100 | Sub-tropical | Jan18-Aug18 | Dry, rainy | 66 000 | 37 | 312 | 31 | 28 (22, 38) |
| **Nepal** | Reuse of population survey<sup>47</sup> | Proposed risk factors | Nepal | 67 | Sub-tropical to Arctic | 2016-2018 | All year | 29 000 000 | 92 | 8 916 | 37 | 41 (33, 50) |
| **Nicaragua** | 1. CKD focused study<sup>3</sup> | Reported high CKDu area | Chinandega (banana/sugarcane) | 0 | Tropical | Jul07-Oct07 | Rainy | 384 | 86 | 331 | 47 | 34 (26, 44) |
|  |  |  | Chinandega (service) | 0 | Tropical | Jul07-Oct07 | Rainy | 177 | 79 | 140 | 36 | 32 (25, 43) |
|  |  |  | Leon (coffee) | 0 | Tropical | Jul07-Oct07 | Rainy | 92 | 84 | 77 | 52 | 36 (27, 46) |
|  |  |  | Leon (fishing) | 0 | Tropical | Jul07-Oct07 | Rainy | 216 | 77 | 166 | 46 | 32 (25, 44) |
|  |  |  | Leon (mining) | 0 | Tropical | Jul07-Oct07 | Rainy | 445 | 86 | 382 | 41 | 33 (26, 43) |
|  | 2. CKD focused study<sup>48</sup> | Reported high CKDu area | Leon municipality | 70 | Tropical | Jun14-Sep14 | Rainy | 204 000 | 97 | 1 672 | 39 | 37 (28, 48) |
| **Peru** | CKD focused study<sup>49</sup> | DEGREE registered | Tumbes | 94 | Arid and Sub-tropical | Nov17-May18 | Spring, summer, autumn | 224 863 | 83 | 1 238 | 43 | 39 (30, 49) |
| **Sri Lanka** | Anuradhapura District - CKD focused study<sup>6</sup> | DEGREE registered | Halambagaswewa, Rambewa | 0 | Tropical | Mar17-May17 | Dry | 1 188 | 90 | 739 | 33 | 41 (33, 49) |
|  |  |  | Lolugaswewa, Medawachchiya | 0 | Tropical | Mar17-May17 | Dry | 1 262 | 86 | 790 | 28 | 41 (34, 50) |
|  |  |  | Pothana, Mihintale | 0 | Tropical | Mar17-May17 | Dry | 1 391 | 88 | 691 | 28 | 41 (33, 51) |
|  |  |  | Puhudivula, Medawachchiya | 0 | Tropical | Mar17-May17 | Dry | 1 362 | 91 | 798 | 28 | 41 (32, 50) |
|  |  |  | Sangilikandarawa, Rambewa | 0 | Tropical | Mar17-May17 | Dry | 1 228 | 90 | 818 | 33 | 41 (33, 50) |
| **Thailand** | Reuse of population survey<sup>50</sup> | Proposed risk factors | Bangkok | 100 | Tropical | Nov13-Aug14 | Cool, hot, rainy | 6 969 010<sup>g</sup> | 81 | 1 604 | 24 | 48 (40, 55) |
|  |  |  | Central | 46 | Tropical | Nov13-Aug14 | Cool, hot, rainy | 14 424 785<sup>g</sup> | 92 | 2 752 | 41 | 46 (35, 54) |
|  |  |  | North | 35 | Tropical | Nov13-Aug14 | Cool, hot, rainy | 8 638 732<sup>g</sup> | 83 | 2 447 | 45 | 47 (37, 54) |
|  |  |  | North East | 29 | Tropical | Nov13-Aug14 | Cool, hot, rainy | 13 445 305<sup>g</sup> | 80 | 2 315 | 46 | 46 (36, 53) |
|  |  |  | South | 34 | Tropical | Nov13-Aug14 | Cool, hot, rainy | 6 442 937<sup>g</sup> | 73 | 2 019 | 42 | 44 (34, 53) |
| **USA** | NHANES 2017-8 – Reuse of population survey<sup>22</sup> | Reference | USA | 83 | All types | 2017-2018 | All year | 320 842 721 | 49 | 3373 | 47 | 39 (30, 49) |

## Methods

The DEGREE collaboration aims to gain insight into the burden of CKDu by using standard protocols to estimate the prevalence of low eGFR in population-representative surveys; the detailed rationale and methods have previously been published.^19^ Here we use the term CKDu to describe the endemic kidney disease of unknown cause occurring at epidemic levels in geographic clusters (i.e., the disease(s) also termed Mesoamerican nephropathy, Uddanam nephropathy or chronic interstitial nephritis in agricultural communities) rather than all forms of CKD without a diagnosis. Defining CKDu is challenging, both at the individual level, and for epidemiological studies, as there is no gold-standard diagnostic test, and diagnosis currently relies on the exclusion of known causes of kidney disease, with only a small number of cases fully documented as tubulointerstitial disease with a kidney biopsy. For these international comparisons of general population prevalence, we have used a pragmatic definition of an eGFR <60ml/min/1.73m^2^ in the absence of diabetes, hypertension, and heavy proteinuria in the working-age population as a surrogate indicator of CKDu burden.

Another important consideration when conducting international comparisons of eGFR is analytical variability in laboratory assays. In this analysis, all studies used standardised isotopic dilution mass spectrometry (IDMS) referenced creatinine measurements, which should minimise this problem, although inter-laboratory and time-dependent variation are still present.^20^ Note that we do not have written confirmation for Nicaragua 1, but this was conducted in a Ministry of Health laboratory where IDMS references were being used at that time. Similar quality control methods are not widely used for cystatin C determination. For our studies, the cystatin C measurements for India, Malawi and Peru were all standardised to a central reference laboratory, but the cystatin C data from Kenya were not standardised.

There were 11 studies formally registered with DEGREE that agreed to conduct population surveys using the DEGREE protocol. Of these, 10 provided data for this analysis. In addition, we identified 11 other studies, using methodology compatible with the DEGREE protocol, that had already been conducted in areas with reported high CKDu prevalence or in settings with proposed CKDu risk-factors. The organisers of these other studies were therefore invited to contribute their data to the joint analyses, of whom seven responded positively and provided data. The studies varied both in the size of the sample and the size of the source population, from focused surveys of specific communities to regional or national surveillance projects (details in Table 1). However, all surveyed the general population of the relevant area (most using either simple random sampling or multi-stage cluster random sampling, see supplemental Table S1). Of the 17 collaborating studies, seven provided us with their data in tabular form, whereas 10 provided us with individual level datasets to create the relevant tables (see supplemental Table S1). Additionally, publicly available data from health surveys in England^21^ and the USA^22^ were obtained to provide reference data from high-income countries. Thus, a total of 19 studies were involved in the current analysis, each reporting data from one or more separately sampled areas.

Populations vary in their age-distribution, and to make our country-comparisons fair, the main outcome was the age-standardised prevalence of eGFR<60ml/min/1.73m^2^ (using the WHO global standard population^23^) in those without hypertension, diabetes, and heavy proteinuria (eGFR<60_[absent HT,DM,high ACR]_), for working-age adults, stratified by rural-urban classification (except the USA where this was not available) and sex (details in Supplementary Text 1).

We also calculated the overall prevalence of eGFR<60ml/min/1.73m^2^ without excluding the population with hypertension, diabetes, or heavy proteinuria (eGFR<60), for comparison. Confidence intervals were calculated for all standardised prevalence estimates.

Except where indicated in supplemental Table S1, eGFR was calculated using the creatinine-based CKD-EPI 2009 equation^24^ but without race adjustment; heavy proteinuria was defined by an albumin-to-creatinine ratio (ACR) of >300mg/g or ≥++ when studies used dipstick urinalysis; diabetes was determined by self-report or HbA1c ≥6.5%; and hypertension was determined by self-report, treatment, systolic blood pressure ≥140mmHg, or diastolic blood pressure ≥90mmHg.

To better understand any selection bias impacting the prevalence estimates, we compared the prevalence of eGFR<60 in the whole available sample to those with complete data for hypertension, diabetes, and proteinuria (before making any exclusions).

Similar analyses were completed using secondary outcomes with a cut-off of 90ml/min/1.73m^2^(eGFR<90 and eGFR<90) to help understand the distribution of low to moderate kidney function and whether the patterns follow or differ to that of low eGFR.

The main analysis used eGFR calculated from serum creatinine, but in a subset, data were available to calculate eGFR using serum cystatin C. Within this subset we compared the results from the original creatinine-based equation to the CKD-EPI 2012 cystatin C only equation and combined creatinine and cystatin C equation.^25^ We also calculated Lin’s concordance correlation coefficient on the individual eGFR data to compare the different measurements.

Prevalence estimates of the main outcome (eGFR<60_[absent HT,DM,high ACR]_) were plotted on international maps, categorised into low (<2%), moderate (2-5%) and high (>5%), to enable visualisation of geographical differences.

Finally, we undertook some sensitivity and other supplementary analyses as follows:

1. Where individual-level data were available, we ran a sensitivity analysis using age-dependent cut-offs of eGFR from a 2020 paper by Jonsson.^26^
2. We ran another sensitivity analysis to consider different eGFR equations using serum creatinine, including CKD-EPI 2021 and CKD MDRD, where it was possible to calculate.
3. We looked for any associations between the main prevalence outcome and study/sample characteristics, including response rate, the proportion of males in the sample, and date of the study.

Data were analysed using Stata version 17^27^ and maps were created using the free open-source QGIS software.^28^

## Results

The characteristics of the 19 studies and 43 areas, including study rationale, response rates and the size of representative populations are shown in Table 1 (with location maps in Supplementary Figure S1). Most studies were in tropical regions and LMICs. The studies were undertaken at different times, ranging from 2007 in Leon and Chinandega, Nicaragua to 2023 in Molina, Chile. The proportion of men in each sample varied from 24%-53% with a median of 43%. The median age varied from 28 years (IQR=[22, 38]) in Lilongwe, Malawi to 53 [48, 57] in Molina, Chile. Stratifying by sex and using age-standardisation mitigates these differences to allow for valid comparisons.

Response rates were mainly high (above 75% and up to 98%), with the exceptions of the high-income reference datasets (England 59%, USA 49%) and the Ecuador (61%), Guatemala (58% and 69%) and Malawi (66% and 37%) studies plus one area of Thailand (South 73%).

Overall, 2015 (3.2%) participants were missing data on hypertension, diabetes, or proteinuria, used in the exclusions, leaving a total sample size of 60 964. The study with the most missing data on these factors was Nepal, where 874 participants (8.9%) had missing data. For all areas considered, estimates of the prevalence of eGFR<60 in the total sample were very similar to those in the sample with complete data (Table S2).

There were 22 255 (36.5%) participants identified as having one or more conditions of hypertension, diabetes, and heavy proteinuria, leaving a sample size of 38 709 for the restricted analyses. The proportion of participants with these conditions varied greatly by area, ranging from about 16% in two Kenyan areas to over 50% in four areas of India. Some of this difference could be explained by the age structure of the samples (as this is before age-standardisation) (Tables 1 and 2).

The age-standardised prevalence estimates of eGFR<60_[absent HT,DM,high ACR]_ stratified by area, sex, and rural-urban classification are shown in Table 2 and Figures 1-4. For men, standardised prevalence estimates of eGFR<60_[absent HT,DM,high ACR]_ were highest in rural areas of Uddanam, India (up to 13.7%, 95% confidence interval (CI) [4.8%, 22.6%]) and areas in northwest Nicaragua (up to 13.6% [6.3%, 20.9%]). Of the other areas considered, prevalence in rural males was low (<2%) in Nepal and some other areas of India and in all areas outside of Central America and South Asia; including Kenya, Peru, Chile, Malawi, and Thailand. High prevalence (>5%) in men was generally only seen in rural areas, but there was one high prevalence urban area in Leon, Nicaragua, and moderate prevalence in Lilongwe, Malawi. There was one low prevalence (<2%) rural area in Nicaragua that was included because residents mainly worked in the service sector. As expected, the prevalence of eGFR<60_[absent HT,DM,high ACR]_ was low in the USA, England, and Italy.

**Figure 1:**
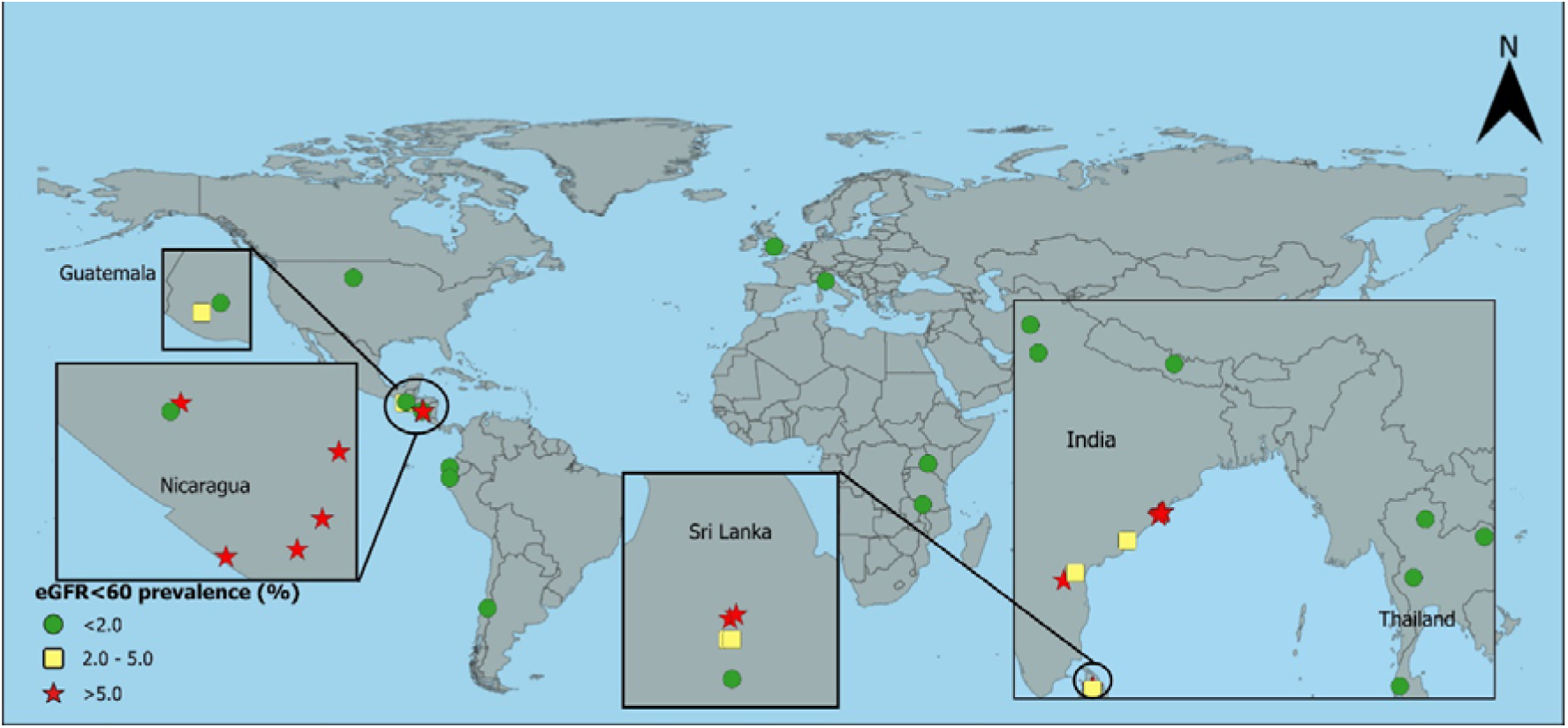
Age-standardised prevalence of creatinine-based eGFR<60 in rural^a^ men without hypertension, diabetes, or heavy proteinuria ^a^USA includes rural and urban together; eGFR=estimated glomerular filtration rate in ml/min/1.73m^2^

**Figure 2:**
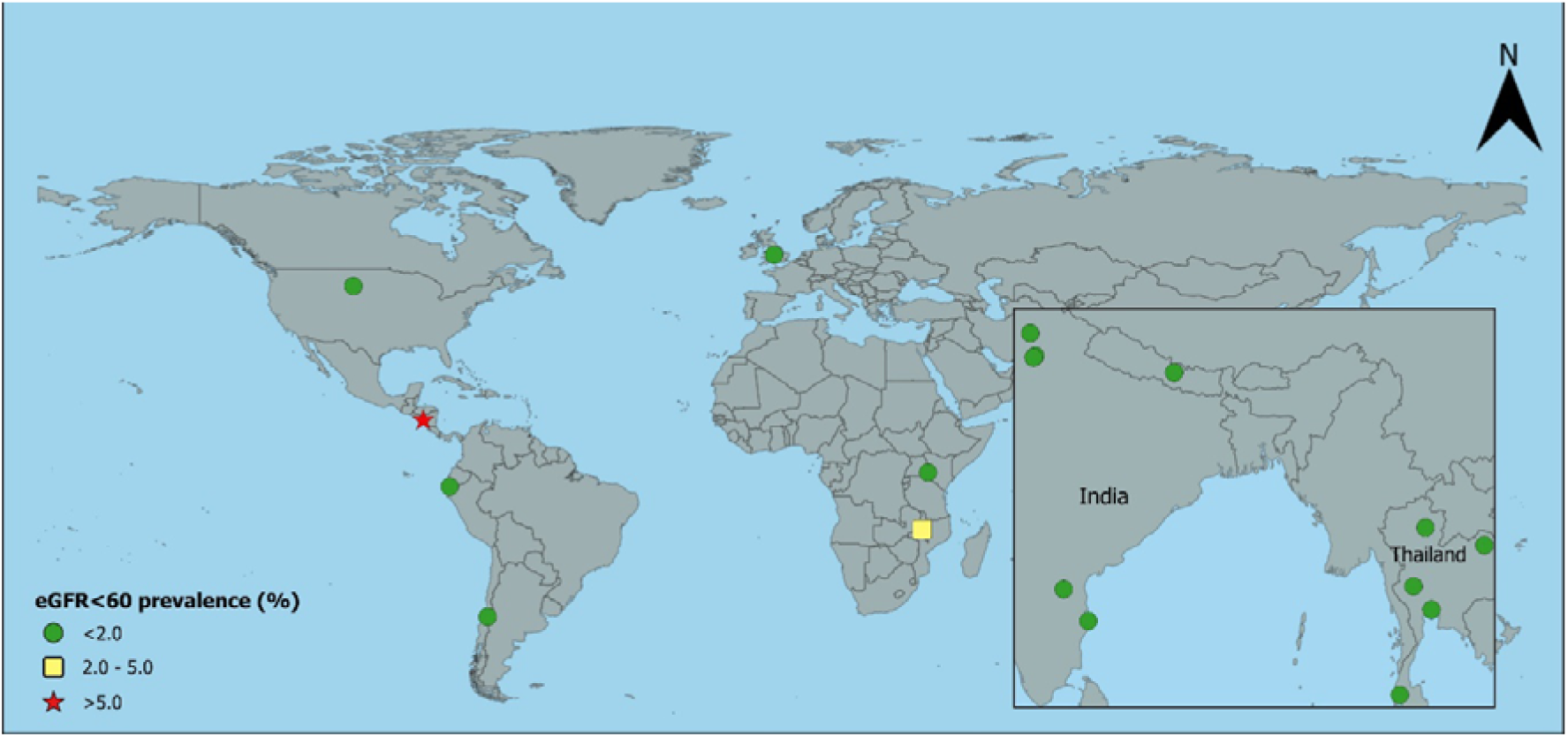
Age-standardised prevalence of creatinine-based eGFR<60 in urban^a^ men without hypertension, diabetes, or heavy proteinuria ^a^USA includes rural and urban together; eGFR=estimated glomerular filtration rate in ml/min/1.73m^2^

**Figure 3:**
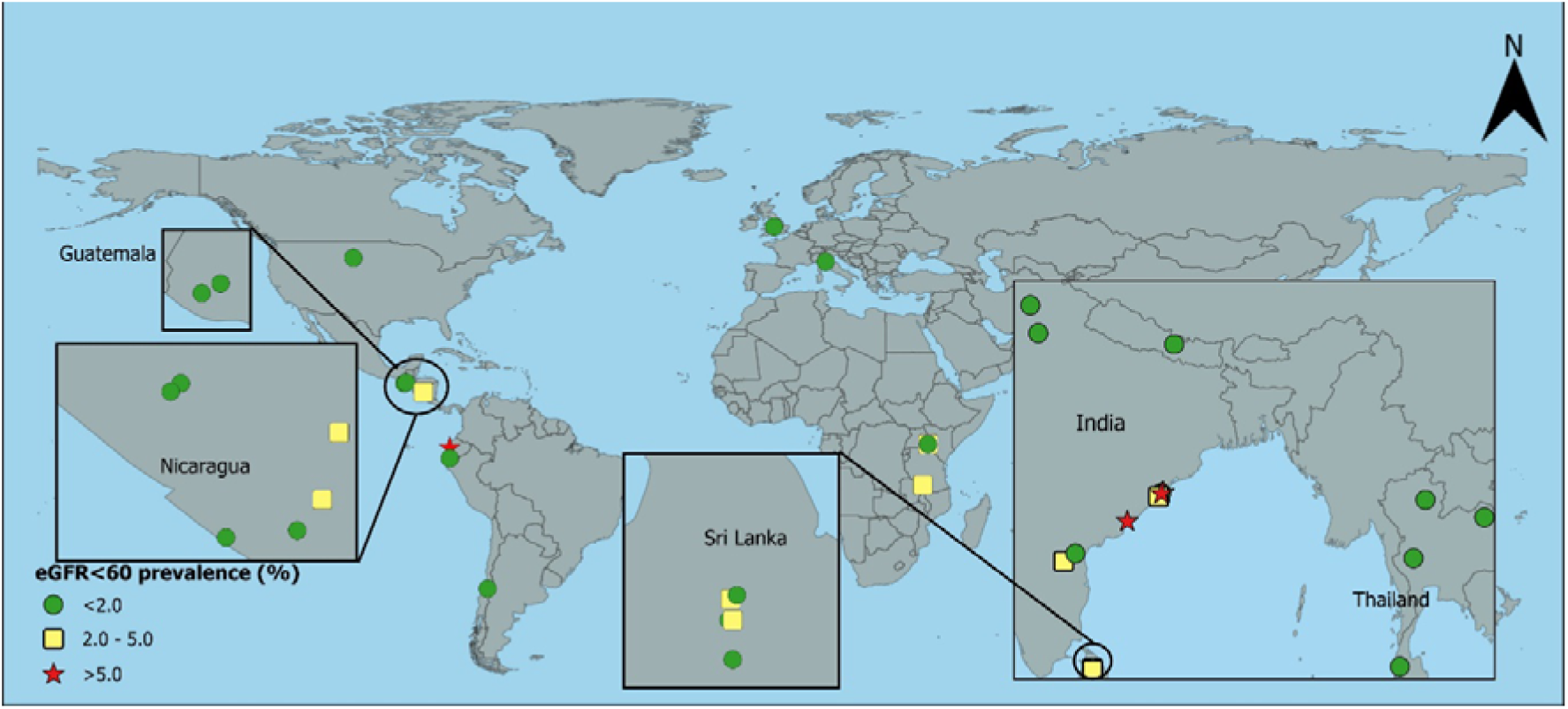
Age-standardised prevalence of creatinine-based eGFR<60 in rural^a^ women without hypertension, diabetes, or heavy proteinuria ^a^USA includes rural and urban together; eGFR=estimated glomerular filtration rate in ml/min/1.73m^2^

**Figure 4:**
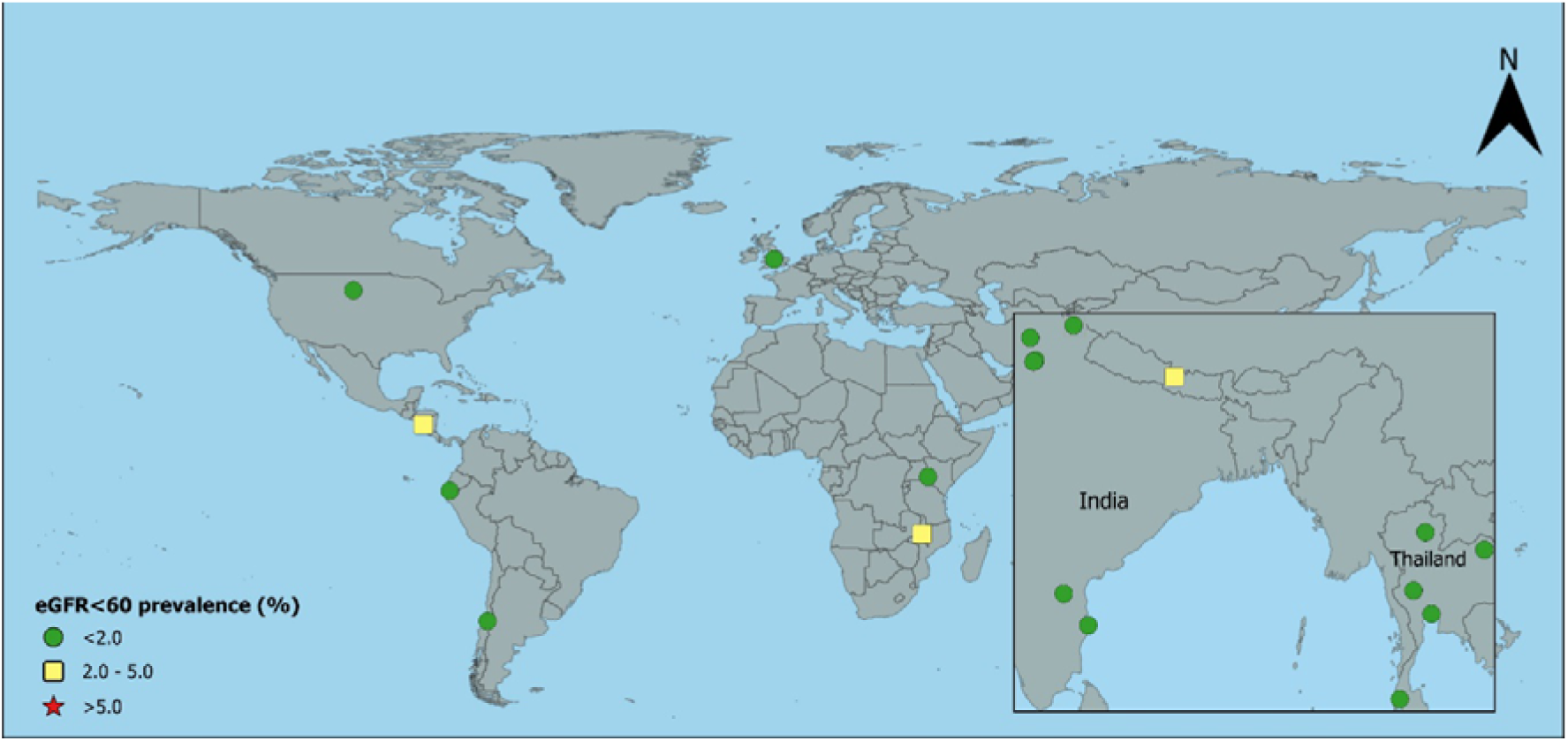
Age-standardised prevalence of creatinine-based eGFR<60 in urban^a^ women without hypertension, diabetes, or heavy proteinuria ^a^USA includes rural and urban together; eGFR=estimated glomerular filtration rate in ml/min/1.73m^2^

**Figure 5:**
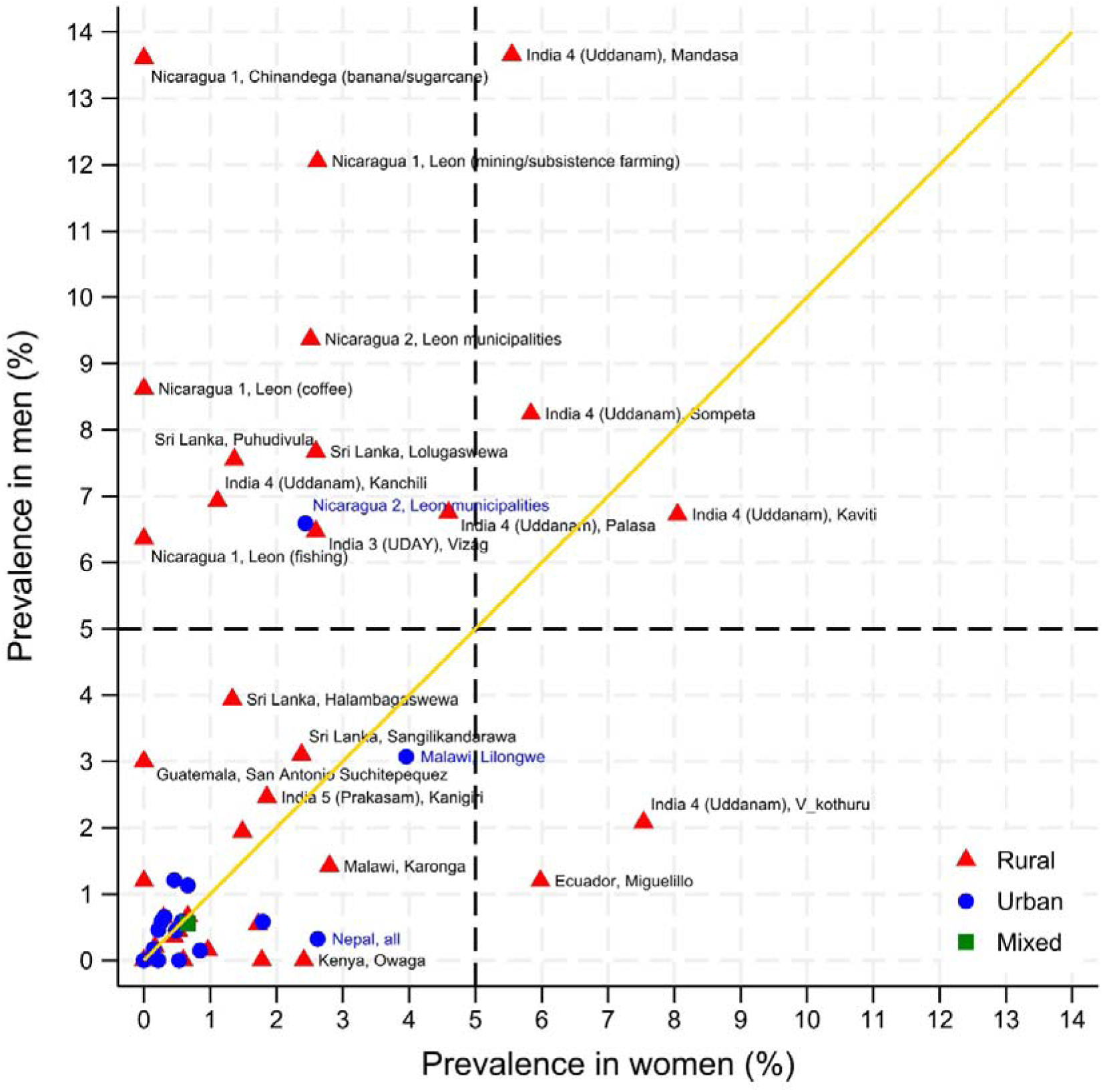
Age-standardised prevalence of eGFR<60 by sex in population without hypertension, diabetes mellitus, or heavy proteinuria eGFR=estimated glomerular filtration rate in ml/min/1.73m^2^

**Table 2:** Age-standardised prevalence of eGFR<60 ml/min/1.73m by sex, for ages 18-60.

| Centre | Area | Rural / Urban | Sample with complete data |  |  |  | Sample of people without hypertension, diabetes, or heavy proteinuria |  |  |  |
| --- | --- | --- | --- | --- | --- | --- | --- | --- | --- | --- |
|  |  |  | Men |  | Women |  | Men |  | Women |  |
|  |  |  | n | eGFR<60<sup>a</sup><br>% (95% CI) | n | eGFR<60<sup>a</sup><br>% (95% CI) | n | eGFR<60<sup>a</sup><br>% (95% CI) | n | eGFR<60<sup>a</sup><br>% (95% CI) |
| Chile<sup>b</sup> | Molina | Rural | 39 | 0.0 (N/A) | 17 | 0.0 (N/A) | 16 | 0.0 (N/A) | 6 | 0.0 (N/A) |
| Chile<sup>b</sup> | Molina | Urban | 156 | 0.0 (N/A) | 264 | 1.6 (0.1, 3.1) | 66 | 0.0 (N/A) | 137 | 0.5 (0, 1.6) |
| Ecuador | Miguelillo | Rural | 312 | 2.2 (0.7, 3.8) | 442 | 6.4 (4.3, 8.5) | 180 | 1.2 (0, 2.8) | 235 | 6.0 (2.2, 9.7) |
| England | all | Rural | 161 | 0.4 (0, 1.0) | 223 | 2.2 (0.8, 3.7) | 98 | 0.0 (N/A) | 169 | 1.8 (0.4, 3.2) |
| England | all | Urban | 744 | 0.5 (0.1, 0.9) | 1007 | 1.0 (0.6, 1.5) | 515 | 0.1 (0, 0.4) | 759 | 0.8 (0.3, 1.4) |
| Guatemala | San Antonio Suchitepequez | Rural | 115 | 3.1 (0, 6.3) | 221 | 0.9 (0, 2.2) | 86 | 3.0 (0, 7.0) | 171 | 0.0 (N/A) |
| Guatemala | Tecpan | Rural | 109 | 0.9 (0, 2.6) | 209 | 0.0 (N/A) | 83 | 0.0 (N/A) | 152 | 0.0 (N/A) |
| India 1 (CARRS) | Chennai | Urban | 2333 | 0.9 (0.5, 1.3) | 3033 | 0.6 (0.3, 0.9) | 1161 | 0.5 (0.0, 0.9) | 1915 | 0.2 (0, 0.5) |
| India 1 (CARRS) | Delhi | Urban | 1733 | 1.0 (0.6, 1.5) | 1831 | 1.2 (0.8, 1.6) | 770 | 0.6 (0.1, 1.1) | 935 | 0.6 (0.0, 1.1) |
| India 2 (ICMR) | Delhi | Urban | 837 | 1.2 (0.6, 1.9) | 1051 | 2.3 (1.5, 3.1) | 399 | 0.6 (0, 1.3) | 571 | 1.8 (0.6, 3.0) |
| India 2 (ICMR) | Faridabad | Rural | 629 | 1.6 (0.8, 2.4) | 784 | 1.6 (0.9, 2.4) | 380 | 1.9 (0.8, 3.1) | 520 | 1.5 (0.5, 2.5) |
| India 3 (UDAY) | Sonipat | Rural | 768 | 0.6 (0.2, 1.1) | 1136 | 0.6 (0.2, 1.0) | 530 | 0.4 (0.0, 0.8) | 847 | 0.5 (0.1, 0.9) |
| India 3 (UDAY) | Sonipat | Urban | 1038 | 0.9 (0.4, 1.3) | 1184 | 0.6 (0.2, 0.9) | 586 | 0.6 (0.1, 1.1) | 768 | 0.3 (0, 0.6) |
| India 3 (UDAY) | Vizag | Rural | 934 | 6.7 (2.9, 10.4) | 1242 | 3.1 (2.2, 4.1) | 696 | 6.5 (1.9, 11.0) | 933 | 2.6 (1.5, 3.7) |
| India 3 (UDAY) | Vizag | Urban | 903 | 1.2 (0.6, 1.8) | 1130 | 0.7 (0.3, 1.1) | 469 | 0.4 (0, 1.1) | 692 | 0.5 (0, 1.1) |
| India 4 (Uddanam) | Kanchili | Rural | 148 | 4.8 (1.1, 8.6) | 169 | 7.2 (3.2, 11.3) | 71 | 6.9 (1.0, 12.9) | 83 | 1.1 (0, 3.2) |
| India 4 (Uddanam) | Kaviti | Rural | 102 | 12.2 (7.7, 16.7) | 110 | 8.0 (3.5, 12.4) | 52 | 6.7 (1.3, 12.2) | 65 | 8.0 (2.0, 14.1) |
| India 4 (Uddanam) | Mandasa | Rural | 95 | 18.4 (10.7, 26.1) | 105 | 10.6 (6.0, 15.1) | 57 | 13.7 (4.8, 22.6) | 54 | 5.6 (0.4, 10.7) |
| India 4 (Uddanam) | Palasa | Rural | 186 | 11.2 (5.9, 16.5) | 176 | 11.0 (6.1, 15.9) | 84 | 6.8 (0.1, 13.4) | 81 | 4.6 (0.5, 8.7) |
| India 4 (Uddanam) | Sompeta | Rural | 202 | 9.2 (4.9, 13.4) | 241 | 2.7 (0.9, 4.5) | 99 | 8.2 (2.2, 14.3) | 127 | 5.8 (1.1, 10.6) |
| India 4 (Uddanam) | V_kothuru | Rural | 250 | 5.3 (3.0, 7.5) | 281 | 4.9 (2.9, 7.0) | 101 | 2.1 (0, 4.8) | 121 | 7.5 (3.2, 11.9) |
| India 5 (Prakasam) | Kanigiri | Rural | 420 | 5.3 (3.4, 7.2) | 632 | 3.4 (2.0, 4.7) | 221 | 2.5 (0.3, 4.6) | 432 | 1.9 (0.7, 3.1) |
| Italy | Barga | Rural | 128 | 0.9 (0, 2.3) | 173 | 0.7 (0, 1.7) | 73 | 1.2 (0, 3.5) | 149 | 0.0 (N/A) |
| Kenya | Muhoroni East | Urban | 138 | 0.0 (N/A) | 122 | 0.9 (0, 2.7) | 113 | 0.0 (N/A) | 104 | 0.0 (N/A) |
| Kenya | Owaga | Rural | 113 | 0.0 (N/A) | 129 | 2.2 (0, 4.8) | 88 | 0.0 (N/A) | 98 | 2.4 (0, 5.8) |
| Kenya | Tonde | Rural | 114 | 0.0 (N/A) | 119 | 0.0 (N/A) | 94 | 0.0 (N/A) | 100 | 0.0 (N/A) |
| Malawi | Karonga | Rural | 271 | 3.0 (0.7, 5.4) | 375 | 3.0 (1.0, 5.1) | 214 | 1.4 (0, 3.5) | 309 | 2.8 (0.3, 5.3) |
| Malawi | Lilongwe | Urban | 96 | 2.2 (0, 6.2) | 216 | 4.9 (1.4, 8.4) | 74 | 3.1 (0, 8.6) | 159 | 4.0 (0.7, 7.2) |
| Nepal | all | Rural | 1690 | 0.9 (0.6, 1.3) | 2790 | 2.1 (1.6, 2.6) | 1002 | 0.5 (0.1, 1.0) | 1981 | 1.7 (1.1, 2.3) |
| Nepal | all | Urban | 1602 | 0.8 (0.4, 1.1) | 2834 | 3.0 (2.4, 3.6) | 822 | 0.3 (0.0, 0.7) | 1777 | 2.6 (1.9, 3.4) |
| Nicaragua 1 | Chinandega (banana/sugarcane) | Rural | 155 | 19.0 (12.5, 25.5) | 176 | 3.5 (0.6, 6.4) | 104 | 13.6 (6.3, 20.9) | 111 | 0.0 (N/A) |
| Nicaragua 1 | Chinandega (service) | Rural | 50 | 0.0 (N/A) | 90 | 0.0 (N/A) | 34 | 0.0 (N/A) | 58 | 0.0 (N/A) |
| Nicaragua 1 | Leon (coffee) | Rural | 40 | 6.3 (0, 14.4) | 37 | 0.0 (N/A) | 30 | 8.6 (0, 19.1) | 26 | 0.0 (N/A) |
| Nicaragua 1 | Leon (fishing) | Rural | 76 | 10.2 (3.2, 17.3) | 90 | 2.1 (0, 5.9) | 55 | 6.4 (0, 13.0) | 73 | 0.0 (N/A) |
| Nicaragua 1 | Leon (mining/subsistence farming) | Rural | 158 | 16.2 (10.4, 22.0) | 224 | 4.8 (1.7, 7.9) | 106 | 12.1 (4.5, 19.6) | 144 | 2.6 (0, 6.6) |
| Nicaragua 2 | Leon municipalities | Rural | 247 | 15.3 (10.9, 19.7) | 329 | 3.1 (1.2, 5.0) | 145 | 9.4 (4.4, 14.3) | 211 | 2.5 (0, 5.4) |
| Nicaragua 2 | Leon municipalities | Urban | 400 | 10.0 (7.2, 12.7) | 696 | 3.6 (2.3, 4.8) | 256 | 6.6 (3.0, 10.1) | 436 | 2.4 (1.0, 3.9) |
| Peru | Tumbes | Rural | 278 | 0.5 (0, 1.3) | 344 | 0.3 (0, 0.9) | 210 | 0.0 (N/A) | 285 | 0.6 (0, 1.7) |
| Peru | Tumbes | Urban | 257 | 0.5 (0, 1.2) | 359 | 0.5 (0, 1.1) | 186 | 0.0 (N/A) | 305 | 0.0 (N/A) |
| Sri Lanka | Halambagaswewa | Rural | 242 | 8.3 (5.5, 11.0) | 497 | 3.8 (2.2, 5.3) | 136 | 3.9 (1.1, 6.8) | 336 | 1.3 (0.1, 2.6) |
| Sri Lanka | Lolugaswewa | Rural | 221 | 5.8 (3.5, 8.1) | 569 | 4.5 (3.1, 5.9) | 138 | 7.7 (3.9, 11.5) | 372 | 2.6 (1.0, 4.2) |
| Sri Lanka | Pothana | Rural | 194 | 5.0 (2.6, 7.3) | 497 | 1.7 (0.7, 2.6) | 115 | 0.7 (0, 1.9) | 330 | 0.7 (0, 1.6) |
| Sri Lanka | Puhudivula | Rural | 222 | 7.7 (5.2, 10.2) | 576 | 3.6 (2.3, 5.0) | 112 | 7.6 (3.8, 11.3) | 378 | 1.4 (0.2, 2.6) |
| Sri Lanka | Sangilikandarawa | Rural | 270 | 6.1 (3.8, 8.4) | 548 | 2.9 (1.6, 4.3) | 157 | 3.1 (0.6, 5.6) | 346 | 2.4 (0.8, 3.9) |
| Thailand | Bangkok | Urban | 381 | 1.1 (0.1, 2.2) | 1223 | 0.7 (0.0, 1.3) | 233 | 1.2 (0, 2.7) | 943 | 0.5 (0, 1.1) |
| Thailand | Central | Rural | 606 | 0.9 (0.3, 1.4) | 795 | 1.1 (0.4, 1.9) | 424 | 0.2 (0, 0.4) | 576 | 1.0 (0.1, 1.8) |
| Thailand | Central | Urban | 519 | 0.6 (0.1, 1.1) | 832 | 0.4 (0.1, 0.7) | 336 | 0.0 (N/A) | 587 | 0.2 (0, 0.5) |
| Thailand | North | Rural | 668 | 1.1 (0.6, 1.7) | 730 | 1.0 (0.1, 1.8) | 407 | 0.4 (0, 0.9) | 498 | 0.5 (0, 1.3) |
| Thailand | North | Urban | 445 | 1.1 (0, 2.3) | 604 | 1.0 (0.4, 1.6) | 267 | 0.7 (0, 1.9) | 409 | 0.3 (0, 0.7) |
| Thailand | North East | Rural | 577 | 1.3 (0.5, 2.1) | 637 | 0.8 (0.3, 1.3) | 428 | 0.6 (0.1, 1.2) | 479 | 0.3 (0, 0.7) |
| Thailand | North East | Urban | 492 | 0.7 (0, 1.4) | 609 | 0.6 (0.2, 1.0) | 351 | 0.2 (0, 0.5) | 450 | 0.1 (0, 0.4) |
| Thailand | South | Rural | 549 | 0.5 (0.1, 1.0) | 678 | 0.2 (0, 0.5) | 366 | 0.2 (0, 0.6) | 493 | 0.2 (0, 0.5) |
| Thailand | South | Urban | 303 | 0.7 (0.0, 1.4) | 489 | 1.1 (0.3, 2.0) | 193 | 1.1 (0, 2.4) | 356 | 0.7 (0, 1.4) |
| USA | all | All | 1586 | 2.1 (1.4, 2.8) | 1787 | 1.3 (0.8, 1.8) | 925 | 0.6 (0.1, 1.0) | 1143 | 0.7 (0.2, 1.2) |

<sup>a</sup> age-standardised prevalence using WHO global population age weights; <sup>b</sup> only 41-60 years included; eGFR=creatinine-based estimated glomerular filtration rate; CI=confidence interval using normal approximation; N/A=no CI available due to zero estimate

For women, the prevalence of eGFR<60_[absent HT,DM,high ACR]_ was generally low, except rural women had an 8.0% [2.0%, 14.1%] prevalence in one area of Uddanam and 6.0% [2.2%, 9.7%] in Ecuador. There was a moderately high prevalence (2-5%) in women in Malawi and urban women in Nepal (Table 2 and Figures 1 and 2).

Standardised prevalence of eGFR<60 (without exclusions) was generally higher than the standardised prevalence of eGFR<60_[absent HT,DM,high ACR]_ as expected, but followed a similar pattern, being highest in rural Uddanam, India (men up to 18.4%, women up to 11.0%) and rural men in Nicaragua (up to 19.0%) (Table 2).

When considering low-moderate eGFR values (eGFR<90_[absent HT,DM,high ACR]_) there was great variability of prevalence and much higher prevalences in some areas, even those without a high prevalence of eGFR<60_[absent HT,DM,high ACR]_ such as England (Supplementary Table S3).

Concordance between eGFR measurements in individuals calculated using creatinine alone compared to cystatin C alone and both creatinine and cystatin C can be seen in Supplementary Table S4. The standardised prevalence of eGFR<60_[absent HT,DM,high ACR]_ using cystatin C was substantially higher in Sonipat and Vizag, India compared to using creatinine in both men and women (from 12.1-21.3% versus 0.0-6.7%). The equation using both creatinine and cystatin also gave higher prevalence but at a much closer level (0.4-10.1%). In two areas of Kenya, there was zero prevalence with the creatinine equation and the creatinine-cystatin equation, but prevalences of 10.4% and 14.3% in women and 3.6% and 0% in men using cystatin alone, although numbers with cystatin C measures were small. Estimates of the prevalence of eGFR<60_[absent HT,DM,high ACR]_ did not differ substantially by measure in Peru, and were lower in Malawi and England when using cystatin C. Similar patterns were seen for eGFR<60 (without exclusions). (Table 3 and Supplementary Table S5)

**Table 3:** Age-standardised prevalence rates of creatinine- and cystatin C-based eGFR<60 ml/min/1.73m in people without hypertension, diabetes, or heavy proteinuria, by sex, for ages 18-60 years with both creatinine and cystatin C measurements available.

| Centre | Area | Rural / Urban | Men |  |  |  | Women |  |  |  |
| --- | --- | --- | --- | --- | --- | --- | --- | --- | --- | --- |
|  |  |  | n | CKD-EPI 2009<sub>creat</sub><br>% (95% CI) | CHKD-EPI 2012<sub>cys</sub><br>% (95% CI) | CKD-EPI 2012<sub>creat-cys</sub><br>% (95% CI) | n | CKD-EPI 2009<sub>creat</sub><br>% (95% CI) | CHKD-EPI 2012<sub>cys</sub><br>% (95% CI) | CKD-EPI 2012<sub>creat-cys</sub><br>% (95% CI) |
| England | all | Rural | 98 | 0.0 (N/A)<sup>b</sup> | 0.5 (0, 1.4) | -<sup>c</sup> | 169 | 1.8 (0.4, 3.2)<sup>b</sup> | 0.3 (0, 0.8) | -<sup>c</sup> |
| England | all | Urban | 515 | 0.1 (0, 0.4)<sup>b</sup> | 0.8 (0.1, 1.4) | -<sup>c</sup> | 759 | 0.8 (0.3, 1.4)<sup>b</sup> | 0.4 (0, 0.7) | -<sup>c</sup> |
| India 3 (UDAY) | Sonipat | Rural | 177 | 0.3 (0, 0.9) | 14.3 (10.5, 18.0) | 1.7 (0.2, 3.2) | 253 | 0.0 (N/A) | 12.9 (7.3, 18.4) | 0.6 (0, 1.3) |
| India 3 (UDAY) | Sonipat | Urban | 199 | 0.0 (N/A) | 14.1 (10.4, 17.8) | 2.5 (0.7, 4.3) | 273 | 0.0 (N/A) | 14.1 (10.6, 17.6) | 2.8 (0.8, 4.8) |
| India 3 (UDAY) | Vizag | Rural | 269 | 6.7 (0, 15.2) | 21.3 (9.9, 32.6) | 10.1 (1.5, 18.8) | 325 | 2.8 (1.2, 4.5) | 12.1 (8.7, 15.5) | 4.8 (2.7, 7.0) |
| India 3 (UDAY) | Vizag | Urban | 152 | 0.0 (N/A) | 12.5 (3.2, 21.9) | 0.4 (0, 1.2) | 244 | 0.0 (N/A) | 12.8 (8.1, 17.5) | 2.1 (0.1, 4.1) |
| Kenya | Muhoroni East | Urban | 4 | 0.0 (N/A) | 0.0 (N/A) | 0.0 (N/A) | 9 | 0.0 (N/A) | 0.0 (N/A) | 0.0 (N/A) |
| Kenya | Owaga | Rural | 37 | 0.0 (N/A) | 3.6 (0, 10.0) | 0.0 (N/A) | 43 | 0.0 (N/A) | 10.4 (4.8, 16.0) | 0.0 (N/A) |
| Kenya | Tonde | Rural | 19 | 0.0 (N/A) | 0.0 (N/A) | 0.0 (N/A) | 29 | 0.0 (N/A) | 14.3 (3.7, 24.9) | 0.0 (N/A) |
| Malawi | Karonga | Rural | 214 | 1.4 (0, 3.5) | 1.1 (0, 2.6) | 1.8 (0, 4.0) | 309 | 2.8 (0.3, 5.3) | 1.6 (0, 3.8) | 2.3 (0, 4.7) |
| Malawi | Lilongwe | Urban | 74 | 3.1 (0, 8.6) | 0.7 (0, 2.1) | 0.0 (N/A) | 159 | 4.0 (0.7, 7.2) | 0.0 (N/A) | 0.7 (0, 2.1) |
| Peru | Tumbes | Rural | 210 | 0.0 (N/A) | 1.7 (0.1, 3.3) | 0.0 (N/A) | 284 | 0.6 (0, 1.7) | 0.9 (0, 2.0) | 0.0 (N/A) |
| Peru | Tumbes | Urban | 186 | 0.0 (N/A) | 1.5 (0, 3.2) | 0.0 (N/A) | 304 | 0.0 (N/A) | 1.4 (0.1, 2.7) | 0.0 (N/A) |

eGFR=estimated glomerular filtration rate; <sup>a</sup> age-standardised prevalence using WHO global population age weights; <sup>b</sup> includes race adjustment; <sup>c</sup> not available as eGFR values supplied and exact age not available; N/A=no CI available due to zero estimate

Results from the sensitivity analyses can be found in Supplementary Text 2.

## Discussion

Our findings are consistent with, and build upon, previous evidence, suggesting a high general population burden of impaired kidney function in the absence of traditional risk factors in areas of Central America, and South Asia (Sri Lanka and South India). Applying the same definition to reference populations from high-income countries, as expected, demonstrated a low prevalence. A key strength of the approach used is that it only depends on eGFR and is independent of the presence or absence of a kidney disease diagnosis. This is of critical importance as such diagnoses are highly dependent on access to nephrology care, which is extremely limited in many CKDu affected regions, making comparisons that rely on ‘absence of diagnosis’ across regions almost impossible to interpret.

### Summary of findings and comparisons to existing literature

In India, studies with a range of sizes of source populations (from thousands to millions) and conducted both with the specific aim of quantifying CKDu prevalence and as part of non-CKDu focused non-communicable disease surveillance surveys, demonstrated similar patterns. That is of a high general population burden of disease in areas of rural coastal Uddanam, but not in urban areas of South India or urban or rural areas in northern India. Interestingly, in the rural coastal areas of Uddanam, where women may also work in the agricultural sector, the prevalence of eGFR<60_[absent HT,DM,high ACR]_ in women approached or exceeded that in men in some study sites. In the Anuradhapura district of Sri Lanka, we observed a high prevalence among men in two out of five rural communities (with moderate prevalence in another two) with small source populations. However, these communities were specifically selected on the basis of clinical data on CKDu burden, thus, it is impossible to make generalisations as to the burden of disease across the wider district.

In northwest Nicaragua, similar to India, data from both a study with a small source population (of thousands) focused on reported high CKDu communities, and a non-CKDu focused non-communicable disease surveillance survey with a larger source population (hundreds of thousands), demonstrated similar patterns with a high prevalence of eGFR<60_[absent HT,DM,high ACR]_ in men. Unusually, there was also a high prevalence of this outcome in the urban population in the latter study, although it is possible that those living in this urban area may still work in agricultural settings. Unfortunately, we were unable to include data from a national survey conducted in El-Salvador (source populations of millions), but this study used similar definitions and reported a prevalence well above reference levels among rural males.^29^ The single study in Guatemala also showed moderate levels of eGFR<60_[absent HT,DM,high ACR]_ in males living in the lowland population sample but low levels in the high-altitude sample.

Many of the studies were conducted using the DEGREE protocol specifically to explore whether there was a burden of CKDu in areas with similar profiles to those seen in areas reported to be affected by a high disease burden. However, the prevalence of eGFR<60_[absent HT,DM,high ACR]_ in rural males was low in Tumbes, Peru (Pacific Coast Latin America, subtropical climate, agricultural), Manabi Province, Ecuador (Pacific Coast Latin America, tropical, agricultural), Karonga District, Malawi (subtropical, agricultural) and Muhoroni Sub-County, Kenya (subtropical, agricultural [specifically sugarcane]). Interestingly, we did identify high prevalence of eGFR<60_[absent HT,DM,high ACR]_ amongst women in Ecuador, and moderately high amongst urban males and both urban and rural women in Malawi, patterns which are not considered typical of CKDu in Central America and South Asia. The relevance of these latter findings remains unclear.

The Thailand study was a re-analyses of a national population survey with a large source population (millions). Sub-populations with a higher prevalence of individuals meeting the case definition (i.e., localised “hot spots”) may be obscured in the larger sampling frames. However, (i) the prevalence of eGFR<60_[absent HT,DM,high ACR]_ is lower than that in the high-income reference populations, and (ii), the source populations of the individual regions in the Thai study are comparable to other large population surveys (included and not included^29^ in this analysis). This suggests that the general population burden of eGFR<60_[absent HT,DM,high ACR]_, is several fold lower in rural regions of Thailand than in the areas most impacted by CKDu in Central America or India. Another population-based study conducted in Northeastern Thailand (not included in this analysis) reported rates of eGFR<60ml/min of ∼10% (without excluding diabetes, hypertension or heavy proteinuria),^30^ but this was almost entirely driven by participants over 60 years of age, and estimates in the working age population were completely consistent with those reported in the analysis included in the current study. The aggregated data from Nepal was derived from a very large source population, and as reports of possible CKDu are mainly focused on returning migrant workers in this country,^31^ it would likely not be possible to detect a high burden of eGFR<60_[absent HT,DM,high ACR]_ in this group using our approach.

### Limitations

When drawing conclusions about CKDu burden the above findings must be considered in the context of limitations of our approach. The pragmatic definition we have used will of course be prone to misclassification in both directions. For example, the definition we used will lead to the inclusion of a range of non-proteinuric (and moderately-proteinuric, non-hypertensive) chronic kidney diseases of both known (e.g., due to congenital abnormalities, granulomatous, or drug-induced chronic interstitial nephritides) and unknown (but non-CKDu) causes. Furthermore, the absence of confirmatory eGFR measures means a proportion of cases reflect those with acute, rather than chronic, kidney injury. Conversely, some true cases of CKDu were probably excluded, particularly where the disease co-exists with diabetes or hypertension (although this would only have biased the prevalence estimates if the prevalence of CKDu was markedly different in people with these conditions than in those without), or in advanced disease where proteinuria is well described. Given this potential for misclassification, a low disease burden will not be observable using our definitions. Nonetheless, a high general population prevalence of eGFR<60_[absent HT,DM,high ACR]_ clearly identifies regions known to be hotspots of CKDu.

The rationale and scale of the studies included in this analysis varied substantially. Some studies were part of large country-wide non-communicable disease surveys, some were specific to kidney disease but covering smaller areas with typical CKDu population characteristics but without previous reports of a high burden of disease, and others were targeted at specific areas chosen on the expectation that the prevalence was high or low. However, all studies were population representative and although response rates varied, this did not appear to be related to the prevalence of the outcome (Supplementary Table S8).

Similarly, working age men tended to be under-represented in most studies. However, this will not affect the prevalence estimates for this group (i.e., the proportion with low eGFR in the working age men who actually participated), unless specific high-risk subgroups (e.g., men in occupations with high prevalence) are under- or over-represented.

Another important limitation is that the CKD-EPI equation has been reported to substantially overestimate eGFR around the 60ml/min/1.73m^2^ threshold in Indian^32^ and sub-Saharan African^33^ populations, and the validity of the equation is unknown in other groups, such as indigenous Americans. We were able to use cystatin C-based equations which have been shown to be more precise^34^ to address this issue in a number of the studies included in this analysis. This sub-analysis demonstrated an increased proportion meeting the outcome across all regions in the Indian study, although relative patterns of prevalence were preserved. This confirms the challenges surrounding using GFR estimating equations in the Indian population but does not alter the conclusions around the areas most affected by CKDu. This sub-analysis also demonstrated increases in prevalence of the outcome in the Kenyan study, particularly in women, though numbers with cystatin C testing were small, preventing firm conclusions. The cystatin C analysis did not change the major conclusions in the Malawi or Peru studies.

It is important to highlight that although we report substantial variability in age-standardised eGFR<60_[absent HT,DM,high ACR]_ we only aim to describe international patterns in the general population. We identified substantial variability eGFR<60_[absent HT,DM,high ACR]_ even between areas within high prevalence regions, and in both Central America^11,35^ and South Asia^36^ there is evidence supporting an even higher prevalence of CKDu in specific high-risk, i.e., occupational, groups. Therefore, there might be an important burden of CKDu in similar groups located in regions where we have not identified evidence of a high general population prevalence of disease. Only adequately powered, targeted studies in these high-risk populations can address this, and specific studies are therefore needed. Furthermore, other than sex- and urban-rural residence, we have not explored any ecological or individual-level risk factors for eGFR<60_[absent HT,DM,high ACR]_.

Finally, it should be noted that this study was descriptive and intended to identify areas with high burden of disease. It was not intended to identify the causes of CKDu or to explain the observed international patterns. Factors that may affect the international patterns may include differences in exposure to potential risk factors for CKDu (leading hypotheses as to the primary cause of CKDu include occupational heat stress, metal(loid) exposure (particularly in water), and pesticide and particulate matter exposure^1^), differences in the degree of misclassification (e.g., the proportion of non-CKDu causes of low eGFR_[absent HT,DM,high ACR]_) between studies, as well as differences in methodology across the included studies. The patterns we have identified clearly require further research.

## Conclusion

The study findings provide useful estimates of population patterns of low eGFR and are of considerable interest. Taken alongside published evidence, the observations from large surveys and smaller studies support a high general-population burden of CKDu in Central America and Uddanam, India, however there is no evidence for a similar population burden of disease from large surveys in other parts of India or in Thailand. There is also evidence from smaller surveys for a substantial burden of disease, in particular communities in the Anuradhapura district of Sri Lanka, again supporting published evidence. Several other regions surveyed, that have superficially similar characteristics to affected areas (i.e., hot, low-income, agricultural settings) did not demonstrate a prevalence of low eGFR consistent with a high general population burden of CKDu.

## Disclosure

This work was funded by grants from the UK Colt Foundation (CF/02/18) and the UK Medical Research Council (MR/P02386X/1 and MR/V033743/1).

In addition, individual centres received funding as follows: The study in Chile was supported by the Chilean Agency of Research and Development (ANID), FONDECYT grant 1221680, and FONDAP-MAUCO grant 1523A0008; The study in Ecuador was supported by Universidad Internacional del Ecuador grant EDM-INV-04-19; The study in Guatemala was supported by NIH Grant NIH/FIC 5R21TW010831; The Indian study in Uddanam was supported by a Grand Challenge Award from the Indian Council of Medical Research and the State Government of Andhra Pradesh; The Sri Lankan study was supported by the National Science Foundation of Sri Lanka (RPHS/2016/CKDu 07), the Ministry of Health, Nutrition and Indigenous Medicine and the World Health Organization Country Office Sri Lanka. The study in Italy was supported by the European Union’s Horizon 2020 research and innovation programme grant 824484; The Kenyan study was supported by the National Institute for Occupational Safety and Health (NIOSH), Illinois Education and Research Center (ERC) Pilot Project Research Training Grant (T42/OH008672), Environmental and Occupational Health unrestricted global health research funds, Michael Bruton Workplace Safety Foundation Scholarship (2019), UIC Graduate College Award for Graduate Research (2019), Paul Brandt-Rauf Scholarship in Global Health (2018–19), and Donna Farley Global Health Scholarship (2020–21); The UDAY study in India was supported by an unrestricted educational grant from Eli Lilly and Company under the Lilly NCD Partnership Program. The funding agency had no role in the design, conduct or analysis of the study; The Indian study in Prakasam district was supported by the Indian Council of Medical Research, New Delhi; The earlier Nicaraguan study was supported by the Swedish International Agency for Development Cooperation (Sida), through the SAREC project of Bilateral Research Cooperation with UNAN-León and the Sida-Health supported SALTRA; The later Nicaraguan study was supported by a grant from the Dutch National Postcode Lottery provided funding to Solidaridad covering a proportion of the fieldwork costs and by the La Isla Network. The Thai NHES V study was supported by the Bureau of Policy and Strategy, Ministry of Public Health, Thai Health Promotion Foundation, National Health Security Office, Thailand.

In addition to the funding listed above, author disclosures related to this work are as follows: AB is President of Epidemiologia e Prevenzione, a non-profit social enterprise; PJC has received funding from the National Institute for Health and Care Research (NIHR) (grant 134801); BC has received grants from Kidney Research UK, the Colt foundation and AstraZeneca, has participated on the board of the AstraZeneca ORTIZ study, and has leadership roles in ISN Consortium of CKDu Collaborators (i3C), ISN Western European Board, Consortium for the Epidemic of Nephropathy in Central America and Mexico (CENCAM), and Kidney Research UK Grants Commitee; RCR has received grants from AstraZeneca, Novonordisk, Boehringer Ingelheim, Roche, and Chinook, to his institution, fees for consultancy from AstraZeneca, Novonordisk, Boehringer Ingelheim, Chinook, and Bayer, and honoraria payments from AstraZeneca, Novonordisk, Boehringer Ingelheim, Amgen, and Bayer; SC has received a grant from the NIHR (134801); VJ has received consultancy fees from Bayer, AstraZeneca, Visterra, Chinook, Vera, Biocryst, and Otsuka; DN is the UK Kidney Association Director of Informatics research (unrelated to this work); COCG has received grants from the Universitat Oberta de Catalunya (UOC) and Barcelona Institute for Global Health (ISGlobal); PR received funding from the National Institutes of Health (USA) and is Chief Science Officer of the Maya Health Alliance Guatemala, a community based organization; all other authors declare no competing interests.

## Data sharing statement

Data from the reference datasets can be found in the public domain: NHANES at https://www.cdc.gov/nchs/nhanes/index.htm and Health Survey England at https://beta.ukdataservice.ac.uk/datacatalogue/series/series?id=2000021.

Data for the other included studies may be available by contacting the authors of the respective study papers.

## Acknowledgements

We thank all the participating studies, their researchers, and fieldworkers, along with all the participants for enabling this work to be completed.

## Author contributions

N.P. and B.C. conceived and designed the study; N.P., B.C., K.J., J.G., D.N., V.J., A.S., and R.C-R. wrote the study protocol; P.C., P.D., V.J., P.K., S.M., R.R.T., A.B., P.R., M.H.H., A.C., M.D., A.P., A.B-O., C.O’C-G., P.C., N.G., T.R., S.C.W., S.S, C.K., M.G-Q., S.C., A.A. and D.N. collected and contributed data; C.E.R and S.R. were responsible for data cleaning and management; C.E.R. and M.N. analysed and visualised the data; C.E.R, N.P. and B.C. wrote the first draft of the manuscript; all other authors contributed to revisions of the manuscript. Others also contributed to the planning and conduct of the study, and to the revisions of the paper, and are listed below as part of the DEGREE Study Group.

## Appendix

### The DEGREE Study Group

Wichai Aekplakorn, Ramathibodi Hospital, Thailand

Shuchi Anand, Stanford University, USA

Aurora Aragón, Wuqu’ Kawoq Maya Health Alliance, Guatemala

Antonio Bernabe-Ortiz, Universidad Peruana Cayetano Heredia, Peru

Annibale Biggeri, University of Padua, Italy

Emmanuel Burdmann, Sao Paulo University, Brazil

Ben Caplin, University College London, UK

Dolores Catelan, University of Padua, Italy

Pubudu Chulasiri, Ministry of Health, Sri Lanka

Philip Cooper, St George’s University of London, UK; International University of Ecuador, Ecuador

Ricardo Correa-Rotter, National Autonomous University of Mexico, Mexico

Sandra Cortés, Pontifical Catholic University of Chile, Chile

Amelia Crampin, Malawi Epidemiology and Intervention Research Unit (MEIRU), Malawi; University of Glasgow, Scotland

Melissa de Santiago, Johns Hopkins Bloomberg School of Public Health, USA

Meghnath Dhimal, Nepal Health Research Council, Nepal

Chiara Doccioli, University of Florence, Italy

Prabhakaran Dorairaj, Public Health Foundation of India, India

Samuel Dorevitch, University of Illinois Chicago, Kenya

Catterina Ferreccio, Pontifical Catholic University of Chile, Chile

Jason Glaser, La Isla Network, USA

Marvin Gonzalez-Quiroz, The University of Texas Health Science Centre at San Antonio, USA; Wuqu’ Kawoq Maya Health Alliance, Guatemala; University College London, UK

Emily Granadillo, International University of Ecuador, Ecuador

Monsermin Gualan, International University of Ecuador, Ecuador

Balaji Gummidi, George Institute, India

Nalika Gunawardena, WHO, India

Sophie Hamilton, UK Health Security Agency, UK

Michelle Hathaway, University of Illinois Chicago, Kenya

Kristina Jakobsson, University of Gothenburg, Sweden

Prashant Jarhyan, Public Health Foundation of India, India

Vivekanand Jha, George Institute, India

Oomen John, George Institute, India

Richard J Johnson, Colorado University, USA

Prabhdeep Kaur, Indian Institute of Science, India

Chagriya Kitiyakara, Ramathibodi Hospital, Thailand

Pornpimol Kongtip, Mahdiol University, Thailand

Hans Kromhout, Utrecht University, Netherlands

Adeera Levin, University of British Columbia, Canada

Magdalena Madero, Ignacio Chavez Institute, Mexico

Estelle McLean, London School of Hygiene and Tropical Medicine, UK

J. Jaime Miranda, Universidad Peruana Cayetano Heredia, Peru

Joseph Mkandawire, Malawi Epidemiology and Intervention Research Unit (MEIRU), Malawi

Sailesh Mohan, Public Health Foundation of India, India

Sharan Murali, Indian Council of Medical Research - National Institute of Epidemiology (ICMR-NIE), India

Devaki Nair, University College London; Health Service Laboratory, UK

Wisdom Nakanga, Malawi Epidemiology and Intervention Research Unit (MEIRU), Malawi

Dorothea Nitsch, London School of Hygiene and Tropical Medicine, UK

Mary Njoroge, London School of Hygiene and Tropical Medicine, UK

Moffat Nyirenda, Medical Research Council/Uganda Virus Research Institute and London School of Hygiene and Tropical Medicine Uganda Research Unit, Uganda; Malawi Epidemiology and Intervention Research Unit (MEIRU), Malawi

Cristina O’Callaghan-Gordo, Open University of Catalonia, Spain; ISGlobal, Spain; Pompeu Fabra University, Spain; Epidemiology and Public Health area of the Red Biomedical Research Centre (CIBERESP), Spain

Neil Pearce, London School of Hygiene and Tropical Medicine, UK

Anil Poudyal, Nepal Health Research Council, Nepal

Narayan Prasad, Sanjay Gandhi Post Graduate Institute of Medical Sciences, India

Lesliam Quirós-Alcalá, Johns Hopkins Bloomberg School of Public Health, USA

Giuseppe Remuzzi, Istituto Mario Negri, Italy

Steven Robertson, London School of Hygiene and Tropical Medicine, UK

Peter Rohloff, Wuqu’ Kawoq Maya Health Alliance, Guatemala

Natalia Romero-Sandoval, International University of Ecuador, Ecuador

Andrea Ruiz-Alejos, Universidad Peruana Cayetano Heredia, Peru

Charlotte Rutter, London School of Hygiene and Tropical Medicine, UK

Thilanga Ruwanpathirana, Ministry of Health, Sri Lanka

Manikandanesan Sakthivel, Indian Council of Medical Research, India

Rajiv Saran, University of Michigan, USA

Sameera Senanayake, Duke-NUS Medical School, Singapore

Leah Shaw, Wuqu’ Kawoq Maya Health Alliance, Guatemala

Ajay Singh, Harvard University, USA

Liam Smeeth, London School of Hygiene and Tropical Medicine, UK

Camilo Sotomayor, Universidad de Chile, Chile

Ravi Raju Tatapudi, The Apollo University, India

Eva Tuiz, Wuqu’ Kawoq Maya Health Alliance, Guatemala

Nikhil Srinivasapura Venkateshmurthy, Public Health Foundation of India, India

Vidhya Venugopal, Sri Ramachandra Institute of Higher Education and Research, India

SC Wickramasinghe, Ministry of Health, Sri Lanka

