## Supplementary for "International prevalence patterns of low eGFR in adults aged 18-60 without traditional risk factors from population-based cross-sectional studies: a disadvantaged populations eGFR epidemiology (DEGREE) study"

**Supplementary material**

**Supplementary Text 1: Age-standardisation method**

Age-standardised prevalence was calculated by $\sum_{i=1}^{4} ASP_i * w_i$, where $ASP_i$ is the age-specific prevalence in age group i (4 groups including 18-30, 31-40, 41-50, and 51-60 years), and $w_i$ is the weight for each age group i calculated as the global population proportion in age group i divided by the global population proportion in the entire 18-60 years group, using the WHO global standard population.[S1] Note that the Chilean study was an exception, only surveying those over 40, so only age groups 41-50 and 51-60 were included in those calculations.

**Supplementary Text 2: Findings from sensitivity analyses and post-analysis checks**

Using age-adapted cut-offs led to marked increases in prevalence in some areas, particularly Sri Lanka, Nepal and England, whereas other areas did not change (Table S6).

In the studies where it was possible to do the relevant analyses, use of CKD-EPI 2021 did not substantially alter prevalence estimates compared to those obtained using CKD-EPI 2009. In contrast, the MDRD equation showed much higher prevalences in many areas (Table S7).

These sensitivity analyses didn't change any of our major conclusions.

When testing for associations between the main prevalence outcome and study design characteristics, the response rate and the study year were not associated, even when restricting to areas with moderate-high prevalence. There was borderline evidence that a higher proportion of males in the sample was associated with a slightly higher prevalence, but only in areas with moderate-high prevalence (Table S8).

### Supplementary Figures

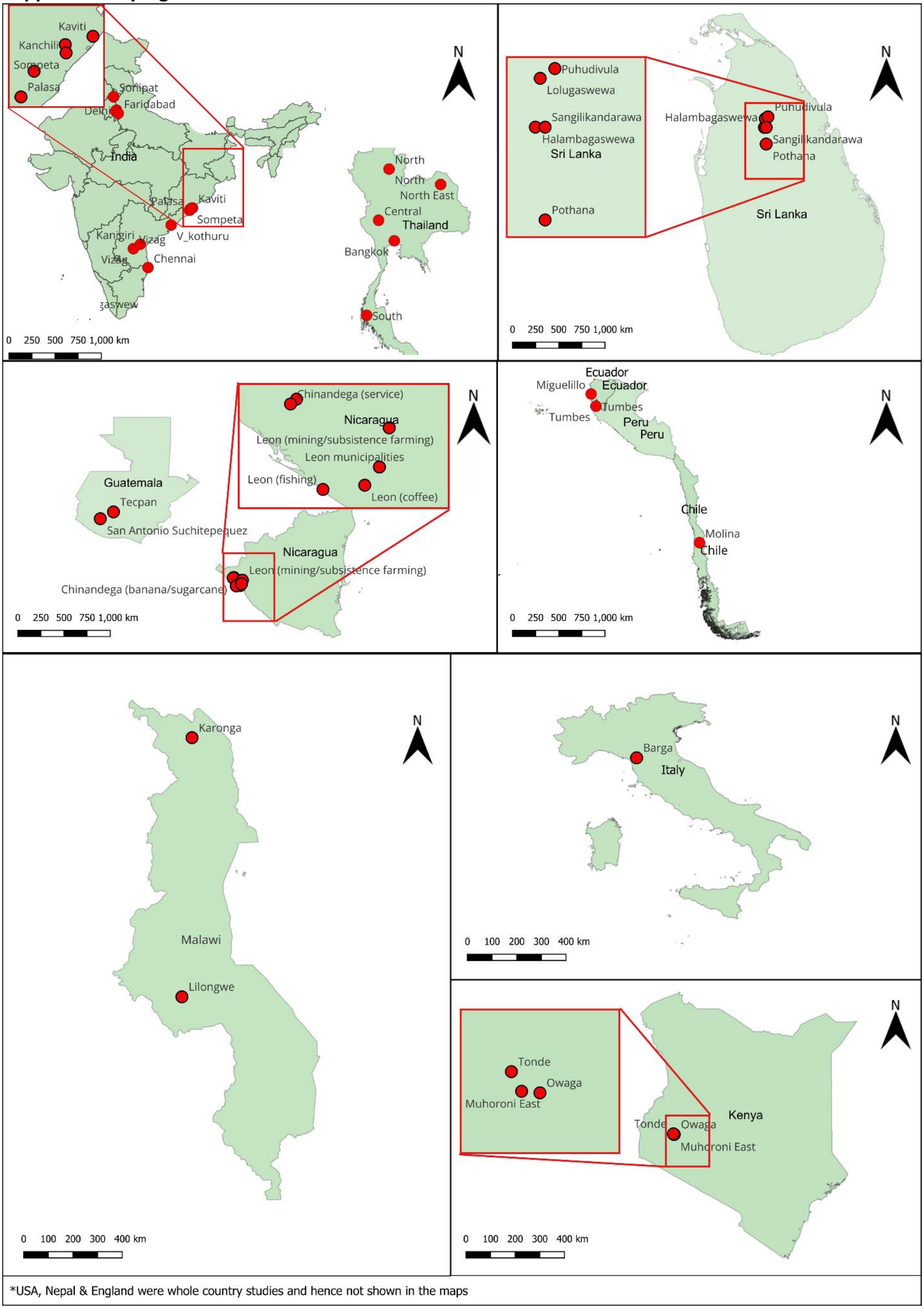


Figure S1: DEGREE centre locations within countries*

### Supplementary tables

Table S1: Definitions of variables and any differences between studies

| Centre | Hypertension definition | Diabetes definition | Heavy proteinuria definition | eGFR calculation in main analysis[a] | Sampling method | Other |
|---|---|---|---|---|---|---|
| Chile | Self-report of diagnosis or mean (of 2) SBP≥140mmHg or DBP≥90mmHg or use of antihypertensive drugs in past 2 weeks | Self-report or glycemia ≥126mg/dL or use of hypoglycemic drugs | urine ACR≥300mg/g | Creatinine: CKD-EPI 2021 | Simple random sampling | Tables provided.<br>Age <=40 excluded. |
| Ecuador | SBP≥140 or DBP≥90 mmHg in two different measurements | HbA1c ≥ 6.5%. | urine dipstick protein ≥+++ | Creatinine: CKD-EPI 2009 (with race) | Convenience sampling. All adults aged 18 years and older living in the communities were invited to participate through community assemblies | Tables provided. |
| England | Self report of diagnosis, or mean (of 3) SBP≥140 mmHg or DBP≥90mmHg | Self report of diagnosis or HbA1c ≥6.5% | urine ACR>300mg/g | Creatinine: CKD-EPI 2009 (with race)<br>Cystatin C: CKD-EPI 2012 | Multi-stage stratified probability sampling | Nationally representative nationwide survey. Age groups are one year out i.e. 18-29, 30-39, 40-49, 50-59.<br>Non-institutionalised individuals only. |
| Guatemala | Self report of diagnosis, or mean (of 3) SBP≥140mmHg or DBP≥90mmHg | Self report of diagnosis or HbA1c ≥6.5% | urine ACR>300mg/g | Creatinine: CKD-EPI 2009 (non race) | Simple random sampling | |
| India 1 (CARRS) | Self-report of diagnosis or mean SBP≥140mmHg or DBP≥90 mmHg | Self report of diagnosis or HbA1c ≥6.5% or fasting glucose ≥126mg/dl | urine ACR>300mg/g | Creatinine: CKD-EPI 2009 (non race) | Multi-stage cluster random sampling | Age <20 years excluded.<br>Pregnant, bedridden and participants who were unable to comprehend the questionnaires due cognitive deficiencies were excluded. |
| India 2 (ICMR) | Self-report of diagnosis or mean SBP≥140mmHg or DBP≥90mmHg | Self report of diagnosis or HbA1c ≥6.5% or fasting glucose ≥126mg/dl | urine ACR>300mg/g | Creatinine: CKD-EPI 2009 (non race) | Multi-stage cluster random sampling for urban area, simple random sampling for rural area | Age <30 years excluded.<br>Pregnant, bedridden and participants who were unable to comprehend the questionnaires due cognitive deficiencies were excluded. |
| India 3 (UDAY) | Self-report of diagnosis or mean SBP≥140mmHg or DBP≥90mmHg | Self report of diagnosis or HbA1c ≥6.5% or fasting glucose ≥126mg/dl | urine ACR>300mg/g | Creatinine: CKD-EPI 2009 (non race)<br>Cystatin C: CKD-EPI 2012 | Multi-stage cluster random sampling | Age <30 years excluded.<br>Pregnant, bedridden and participants who were unable to comprehend the questionnaires due cognitive deficiencies were excluded. |
| India 4 (Uddanam) | Self-report of >5 year hypertension | HbA1c ≥6.5% | urine PCR>150mg/g | Creatinine: CKD-EPI 2012 (non race) | Cluster random sampling | Tables provided. |
| India 5 (Prakasam) | SBP≥140mmHg or DBP≥90mmHg | HbA1c ≥6.5% | urine ACR≥300mg/g | Creatinine: CKD-EPI 2021 (non-race) | Whole population (of selected villages) | Tables provided. |
| Italy | Self-report of diagnosis or SBP≥140mmHg or DBP≥90mmHg | Self-report of diagnosis | urine dipstick protein ≥++ | Creatinine: CKD-EPI 2009 (non race) | Simple random sampling | Stratified by age and sex. |

| | | | | | | |
|---|---|---|---|---|---|---|
| Kenya | Self-report of diagnosis or medication for hypertension, mean SBP≥140mmHg or mean DBP≥90mmHg | Self-report of diagnosis or diabetic medication | urine dipstick protein ≥++ | Creatinine: CKD-EPI 2009 (non race)<br>Cystatin C: CKD-EPI 2012 | Household random sampling | One individual per house selected, with sex predetermined. |
| Malawi | Self-report of diagnosis, or hypertensive medication in last 2 weeks, or mean SBP≥140mmHg or DBP≥90mmHg | Glucose ≥7mmol/L, or self-report previous diagnosis or diabetic medication | urine ACR>300mg/g | Creatinine: CKD-EPI 2009 (non race)<br>Cystatin C: CKD-EPI 2012 | Simple random sampling | |
| Nepal | Diagnosis (verified through medical records provided by participant) or mean SBP≥140mmHg or DBP≥90mmHg | Taking diabetic medication, or fasting glucose ≥126mg/dL or post prandial blood glucose levels ≥200mg/dL | Urine protein>=300 mg/dL | Creatinine: CKD-MDRD | Multi-stage cluster random sampling | Nationally representative nationwide survey. Too frail participants and those who cannot provide written consent were excluded. |
| Nicaragua 1 | Self-report of diagnosis, or SBP≥140mmHg or DBP≥90mmHg | Self-reported diagnosis or glucosuria ≥100mg/dL using a urine stick | urine dipstick protein ≥+++ | Creatinine: CKD-MDRD | Simple random sampling | Tables provided. |
| Nicaragua 2 | Self-report of diagnosis or mean SBP≥140mmHg or DBP≥90mmHg | Self-reported diagnosis or serum glucose ≥110mg/dL | urine dipstick protein ≥+++ | Creatinine: CKD-EPI 2012 (with race) for $eGFR_{creat}$ | Household random sampling | Tables provided.<br>Stratified by urban/rural to match the population rate |
| Peru | Self-report of diagnosis, or mean SBP≥140mmHg or DBP≥90mmHg | Self reported diagnosis, or taking diabetes medication, or fasting glucose ≥126mg/dL | urine dipstick protein ≥++ | Creatinine: CKD-EPI 2009<br>Cystatin C: CKD-EPI 2012 | Simple random sampling | Stratified by urban/rural to 50% and sex. Female recruitment stopped after reaching 60% for each subgroup. |
| Sri Lanka | Self report of taking medication for hypertension, or mean SBP≥140mmHg or DBP≥90mmHg | Self-reported diagnosis, or current diabetic medication, or fasting glucose ≥126mg/dL | urine ACR>300 mg/g | Creatinine: CKD-EPI 2009 | Simple random sampling | Stratified by urban/rural (almost 50% in each group) and by sex. Female recruitment stopped after reaching 60% for each subgroup. |
| Thailand | Self report of taking medication for hypertension, or mean SBP≥140mmHg or DBP≥90mmHg | Use of antihypoglycemic medications, or fasting plasma glucose ≥126mg/dL | urine dipstick protein ≥++ | Creatinine: CKD-EPI 2009 (non race) | Multi-stage cluster random sampling | Tables provided.<br>Stratified, nationally representative, nationwide survey. |
| USA | Self-report of diagnosis, or mean (of 3) SBP≥140mmHg or DBP≥90mmHg | Self report of diagnosis or HbA1c ≥6.5% | urine ACR>300mg/g | Creatinine: CKD-EPI 2009 (non race)$_t$ | Multi-stage cluster random sampling | Nationally representative nationwide survey.<br>Non-institutionalised individuals only.<br>Urban/rural flag unavailable. |

SBP=systolic blood pressue; DBP=diastolic blood pressure; ACR=albumin creatinine ratio; HbA1c=glycated haemoglobin; CKD-EPI=The Chronic Kidney Disease Epidemiology Collaboration; CKD-MDRD=Chronic Kidney Disease Modification of Diet in Renal Disease; [a] All creatinine assays were IDMS (isotopic dilution mass spectrometry) referenced. We do not have written confirmation for Nicaragua 1 but this was conducted in a ministry of health laboratory where IDMS references were being used at the time. Cystatin C measures from India, Malawi and Peru were standardised to a single reference laboratory.

Table S2: Age-standardised prevalence of eGFR<60ml/min/1.73m$^2$ in total sample and complete data sample (including those with diabetes, hypertension and heavy proteinuria), by sex

| Centre | Area | Rural / Urban | Total Sample men | Men with missing data | Total sample prevalence of eGFR<60[a] in men % (95% CI) | Complete case prevalence of eGFR<60[a] in men % (95% CI) | Total Sample women | Women with missing data | Total sample prevalence of eGFR<60[a] in women % (95% CI) | Complete case prevalence of eGFR<60[a] in women % (95% CI) |
|---|---|---|---|---|---|---|---|---|---|---|
| Chile | Molina | Rural | 42 | 3 | 0.0 (N/A) | 0.0 (N/A) | 18 | 1 | 0.0 (N/A) | 0.0 (N/A) |
| Chile | Molina | Urban | 167 | 11 | 0.0 (N/A) | 0.0 (N/A) | 283 | 19 | 1.5 (0.1, 2.9) | 1.6 (0.1, 3.1) |
| Ecuador | Miguelillo | Rural | 312 | 0 | 2.2 (0.7, 3.8) | 2.2 (0.7, 3.8) | 442 | 0 | 6.4 (4.3, 8.5) | 6.4 (4.3, 8.5) |
| England | all | Rural | 167 | 6 | 0.4 (0, 1.0) | 0.4 (0, 1.0) | 233 | 10 | 2.1 (0.7, 3.5) | 2.2 (0.8, 3.7) |
| England | all | Urban | 789 | 45 | 0.6 (0.1, 1.0) | 0.5 (0.1, 0.9) | 1063 | 56 | 1.0 (0.6, 1.5) | 1.0 (0.6, 1.5) |
| Guatemala | San Antonio Suchitepequez | Rural | 119 | 4 | 3.0 (0, 6.0) | 3.1 (0, 6.3) | 224 | 3 | 0.9 (0, 2.2) | 0.9 (0, 2.2) |
| Guatemala | Tecpan | Rural | 116 | 7 | 0.8 (0, 2.4) | 0.9 (0, 2.6) | 236 | 27 | 0.0 (N/A) | 0.0 (N/A) |
| India 1 (CARRS) | Chennai | Urban | 2374 | 41 | 0.9 (0.5, 1.2) | 0.9 (0.5, 1.3) | 3108 | 75 | 0.6 (0.3, 0.9) | 0.6 (0.3, 0.9) |
| India 1 (CARRS) | Delhi | Urban | 1791 | 58 | 1.0 (0.6, 1.5) | 1.0 (0.6, 1.5) | 1890 | 59 | 1.2 (0.7, 1.6) | 1.2 (0.8, 1.6) |
| India 2 (ICMR) | Delhi | Urban | 864 | 27 | 1.2 (0.6, 1.8) | 1.2 (0.6, 1.9) | 1102 | 51 | 2.3 (1.5, 3.1) | 2.3 (1.5, 3.1) |
| India 2 (ICMR) | Faridabad | Rural | 670 | 41 | 1.6 (0.9, 2.4) | 1.6 (0.8, 2.4) | 845 | 61 | 1.6 (0.9, 2.4) | 1.6 (0.9, 2.4) |
| India 3 (UDAY) | Sonipat | Rural | 778 | 10 | 0.6 (0.2, 1.1) | 0.6 (0.2, 1.1) | 1184 | 48 | 0.6 (0.2, 1.0) | 0.6 (0.2, 1.0) |
| India 3 (UDAY) | Sonipat | Urban | 1046 | 8 | 0.9 (0.4, 1.3) | 0.9 (0.4, 1.3) | 1206 | 22 | 0.6 (0.2, 0.9) | 0.6 (0.2, 0.9) |
| India 3 (UDAY) | Vizag | Rural | 1019 | 85 | 6.1 (3.0, 9.1) | 6.7 (2.9, 10.4) | 1349 | 107 | 3.3 (2.2, 4.3) | 3.1 (2.2, 4.1) |
| India 3 (UDAY) | Vizag | Urban | 935 | 32 | 1.2 (0.6, 1.8) | 1.2 (0.6, 1.8) | 1193 | 63 | 0.7 (0.3, 1.1) | 0.7 (0.3, 1.1) |
| India 4 (Uddanam) | Kanchili | Rural | 148 | 0 | 4.8 (1.1, 8.6) | 4.8 (1.1, 8.6) | 169 | 0 | 7.2 (3.2, 11.3) | 7.2 (3.2, 11.3) |
| India 4 (Uddanam) | Kaviti | Rural | 102 | 0 | 12.2 (7.7, 16.7) | 12.2 (7.7, 16.7) | 110 | 0 | 8.0 (3.5, 12.4) | 8.0 (3.5, 12.4) |
| India 4 (Uddanam) | Mandasa | Rural | 95 | 0 | 18.4 (10.7, 26.1) | 18.4 (10.7, 26.1) | 105 | 0 | 10.6 (6.0, 15.1) | 10.6 (6.0, 15.1) |
| India 4 (Uddanam) | Palasa | Rural | 186 | 0 | 11.2 (5.9, 16.5) | 11.2 (5.9, 16.5) | 176 | 0 | 11.0 (6.1, 15.9) | 11.0 (6.1, 15.9) |
| India 4 (Uddanam) | Sompeta | Rural | 202 | 0 | 9.2 (4.9, 13.4) | 9.2 (4.9, 13.4) | 241 | 0 | 2.7 (0.9, 4.5) | 2.7 (0.9, 4.5) |
| India 4 (Uddanam) | V_kothuru | Rural | 250 | 0 | 5.3 (3.0, 7.5) | 5.3 (3.0, 7.5) | 281 | 0 | 4.9 (2.9, 7.0) | 4.9 (2.9, 7.0) |
| India 5 (Prakasam) | Kanigiri | Rural | 420 | 0 | 5.3 (3.4, 7.2) | 5.3 (3.4, 7.2) | 632 | 0 | 3.4 (2.0, 4.7) | 3.4 (2.0, 4.7) |
| Italy | Barga | Rural | 130 | 2 | 0.9 (0, 2.3) | 0.9 (0, 2.3) | 176 | 3 | 0.7 (0, 1.7) | 0.7 (0, 1.7) |
| Kenya | Muhoroni East | Urban | 138 | 0 | 0.0 (N/A) | 0.0 (N/A) | 122 | 0 | 0.9 (0, 2.7) | 0.9 (0, 2.7) |
| Kenya | Owaga | Rural | 138 | 25 | 0.0 (N/A) | 0.0 (N/A) | 151 | 22 | 2.1 (0, 4.5) | 2.2 (0, 4.8) |
| Kenya | Tonde | Rural | 114 | 0 | 0.0 (N/A) | 0.0 (N/A) | 119 | 0 | 0.0 (N/A) | 0.0 (N/A) |

| | | | | | | | | | | |
|---|---|---|---|---|---|---|---|---|---|---|
| Malawi | Karonga | Rural | 286 | 15 | 3.0 (0.6, 5.3) | 3.0 (0.7, 5.4) | 382 | 7 | 3.0 (1.0, 4.9) | 3.0 (1.0, 5.1) |
| Malawi | Lilongwe | Urban | 96 | 0 | 2.2 (0, 6.2) | 2.2 (0, 6.2) | 216 | 0 | 4.9 (1.4, 8.4) | 4.9 (1.4, 8.4) |
| Nepal | all | Rural | 1796 | 106 | 0.9 (0.5, 1.3) | 0.9 (0.6, 1.3) | 3151 | 361 | 2.1 (1.6, 2.6) | 2.1 (1.6, 2.6) |
| Nepal | all | Urban | 1683 | 81 | 0.8 (0.4, 1.1) | 0.8 (0.4, 1.1) | 3160 | 326 | 3.0 (2.4, 3.5) | 3.0 (2.4, 3.6) |
| Nicaragua 1 | Chinandega (banana/sugarcane) | Rural | 155 | 0 | 19.0 (12.5, 25.5) | 19.0 (12.5, 25.5) | 176 | 0 | 3.5 (0.6, 6.4) | 3.5 (0.6, 6.4) |
| Nicaragua 1 | Chinandega (service) | Rural | 50 | 0 | 0.0 (N/A) | 0.0 (N/A) | 90 | 0 | 0.0 (N/A) | 0.0 (N/A) |
| Nicaragua 1 | Leon (coffee) | Rural | 40 | 0 | 6.3 (0, 14.4) | 6.3 (0, 14.4) | 37 | 0 | 0.0 (N/A) | 0.0 (N/A) |
| Nicaragua 1 | Leon (fishing) | Rural | 76 | 0 | 10.2 (3.2, 17.3) | 10.2 (3.2, 17.3) | 90 | 0 | 2.1 (0, 5.9) | 2.1 (0, 5.9) |
| Nicaragua 1 | Leon (mining/ subsistence farming) | Rural | 158 | 0 | 16.2 (10.4, 22.0) | 16.2 (10.4, 22.0) | 224 | 0 | 4.8 (1.7, 7.9) | 4.8 (1.7, 7.9) |
| Nicaragua 2 | Leon municipalities | Rural | 247 | 0 | 15.3 (10.9, 19.7) | 15.3 (10.9, 19.7) | 329 | 0 | 3.1 (1.2, 5.0) | 3.1 (1.2, 5.0) |
| Nicaragua 2 | Leon municipalities | Urban | 400 | 0 | 10.0 (7.2, 12.7) | 10.0 (7.2, 12.7) | 696 | 0 | 3.6 (2.3, 4.8) | 3.6 (2.3, 4.8) |
| Peru | Tumbes | Rural | 281 | 3 | 0.5 (0, 1.3) | 0.5 (0, 1.3) | 349 | 5 | 0.3 (0, 0.9) | 0.3 (0, 0.9) |
| Peru | Tumbes | Urban | 261 | 4 | 0.5 (0, 1.2) | 0.5 (0, 1.2) | 360 | 1 | 0.5 (0, 1.1) | 0.5 (0, 1.1) |
| Sri Lanka | Halambagaswewa | Rural | 246 | 4 | 8.2 (5.5, 10.9) | 8.3 (5.5, 11.0) | 504 | 7 | 3.8 (2.2, 5.3) | 3.8 (2.2, 5.3) |
| Sri Lanka | Lolugaswewa | Rural | 221 | 0 | 5.8 (3.5, 8.1) | 5.8 (3.5, 8.1) | 576 | 7 | 4.5 (3.1, 6.0) | 4.5 (3.1, 5.9) |
| Sri Lanka | Pothana | Rural | 201 | 7 | 4.9 (2.6, 7.2) | 5.0 (2.6, 7.3) | 504 | 7 | 1.6 (0.7, 2.6) | 1.7 (0.7, 2.6) |
| Sri Lanka | Puhudivula | Rural | 224 | 2 | 7.6 (5.2, 10.1) | 7.7 (5.2, 10.2) | 580 | 4 | 3.6 (2.3, 4.9) | 3.6 (2.3, 5.0) |
| Sri Lanka | Sangilikandarawa | Rural | 276 | 6 | 6.2 (3.9, 8.5) | 6.1 (3.8, 8.4) | 558 | 10 | 3.1 (1.8, 4.4) | 2.9 (1.6, 4.3) |
| Thailand | Bangkok | Urban | 381 | 0 | 1.1 (0.1, 2.2) | 1.1 (0.1, 2.2) | 1223 | 0 | 0.7 (0.0, 1.3) | 0.7 (0.0, 1.3) |
| Thailand | Central | Rural | 606 | 0 | 0.9 (0.3, 1.4) | 0.9 (0.3, 1.4) | 795 | 0 | 1.1 (0.4, 1.9) | 1.1 (0.4, 1.9) |
| Thailand | Central | Urban | 519 | 0 | 0.6 (0.1, 1.1) | 0.6 (0.1, 1.1) | 832 | 0 | 0.4 (0.1, 0.7) | 0.4 (0.1, 0.7) |
| Thailand | North | Rural | 668 | 0 | 1.1 (0.6, 1.7) | 1.1 (0.6, 1.7) | 730 | 0 | 1.0 (0.1, 1.8) | 1.0 (0.1, 1.8) |
| Thailand | North | Urban | 445 | 0 | 1.1 (0, 2.3) | 1.1 (0, 2.3) | 604 | 0 | 1.0 (0.4, 1.6) | 1.0 (0.4, 1.6) |
| Thailand | North East | Rural | 577 | 0 | 1.3 (0.5, 2.1) | 1.3 (0.5, 2.1) | 637 | 0 | 0.8 (0.3, 1.3) | 0.8 (0.3, 1.3) |
| Thailand | North East | Urban | 492 | 0 | 0.7 (0, 1.4) | 0.7 (0, 1.4) | 609 | 0 | 0.6 (0.2, 1.0) | 0.6 (0.2, 1.0) |
| Thailand | South | Rural | 549 | 0 | 0.5 (0.1, 1.0) | 0.5 (0.1, 1.0) | 678 | 0 | 0.2 (0, 0.5) | 0.2 (0, 0.5) |
| Thailand | South | Urban | 303 | 0 | 0.7 (0.0, 1.4) | 0.7 (0.0, 1.4) | 489 | 0 | 1.1 (0.3, 2.0) | 1.1 (0.3, 2.0) |
| USA | all | All | 1597 | 11 | 2.1 (1.4, 2.7) | 2.1 (1.4, 2.8) | 1796 | 9 | 1.3 (0.8, 1.7) | 1.3 (0.8, 1.8) |

eGFR=estimated glomerular filtration rate; CI=confidence interval using normal approximation [a] age-standardised prevalence using WHO global population age weights;

Table S3: Age-standardised prevalence of eGFR<90ml/min/1.73m$^2$ by sex (for age 18-60)

| Centre | Area | Rural / Urban | Sample with complete data | | | | Sample of people without hypertension, diabetes and heavy proteinuria | | | |
|---|---|---|---|---|---|---|---|---|---|---|
| | | | Men | | Women | | Men | | Women | |
| | | | n | eGFR<90[a] % (95% CI) | n | eGFR<90[a] % (95% CI) | n | eGFR<90[a] % (95% CI) | n | eGFR<90[a] % (95% CI) |
| Chile | Molina | Rural | 39 | 36.2 (17.3, 55.2) | 17 | 59.1 (35.7, 82.4) | 16 | 35.9 (9.0, 62.8) | 6 | 100 (NA, NA) |
| Chile | Molina | Urban | 156 | 42.1 (34.2, 50.0) | 264 | 49.0 (42.8, 55.8) | 66 | 47.5 (36.0, 59.0) | 137 | 46.5 (37.7, 55.2) |
| Ecuador | Miguelillo | Rural | 312 | 34.1 (29.2, 39.1) | 442 | 46.3 (41.9, 50.6) | 180 | 33.4 (26.6, 40.3) | 235 | 45.2 (38.6, 51.9) |
| England | all | Rural | 161 | 35.7 (27.7, 43.8) | 223 | 36.1 (29.7, 42.5) | 98 | 35.8 (26.8, 44.7) | 169 | 37.3 (29.9, 44.8) |
| England | all | Urban | 744 | 37.5 (33.9, 41.2) | 1007 | 31.7 (28.9, 34.5) | 515 | 38.1 (34.0, 42.2) | 759 | 32.9 (29.8, 35.9) |
| Guatemala | San Antonio Suchitepequez | Rural | 115 | 6.8 (2.0, 11.6) | 221 | 4.7 (1.9, 7.5) | 86 | 6.3 (0.5, 12.1) | 171 | 3.4 (0.5, 6.3) |
| Guatemala | Tecpan | Rural | 109 | 10.7 (4.9, 16.5) | 209 | 5.7 (2.6, 8.7) | 83 | 9.4 (2.3, 16.4) | 152 | 8.8 (3.6, 13.9) |
| India 1 (CARRS) | Chennai | Urban | 2333 | 8.2 (7.1, 9.2) | 3033 | 5.8 (5.0, 6.7) | 1161 | 6.0 (4.6, 7.5) | 1915 | 5.4 (4.1, 6.7) |
| India 1 (CARRS) | Delhi | Urban | 1733 | 11.4 (10.1, 12.7) | 1831 | 11.9 (10.6, 13.1) | 770 | 10.8 (8.6, 12.9) | 935 | 11.1 (8.9, 13.2) |
| India 2 (ICMR) | Delhi | Urban | 837 | 33.3 (27.8, 38.8) | 1051 | 37.0 (33.9, 40.0) | 399 | 33.2 (25.8, 40.7) | 571 | 36.1 (32.2, 39.9) |
| India 2 (ICMR) | Faridabad | Rural | 629 | 18.6 (16.4, 20.8) | 784 | 18.1 (12.8, 23.4) | 380 | 18.6 (15.6, 21.5) | 520 | 17.2 (11.0, 23.5) |
| India 3 (UDAY) | Sonipat | Rural | 768 | 15.3 (9.1, 21.4) | 1136 | 12.3 (10.0, 14.5) | 530 | 13.7 (7.7, 19.8) | 847 | 12.5 (10.1, 15.0) |
| India 3 (UDAY) | Sonipat | Urban | 1038 | 12.0 (10.5, 13.4) | 1184 | 11.2 (9.2, 13.2) | 586 | 10.9 (9.0, 12.8) | 768 | 9.9 (7.8, 12.0) |
| India 3 (UDAY) | Vizag | Rural | 934 | 30.6 (24.6, 36.6) | 1242 | 20.1 (17.6, 22.6) | 696 | 30.2 (23.4, 37.0) | 933 | 19.1 (16.4, 21.8) |
| India 3 (UDAY) | Vizag | Urban | 903 | 28.3 (23.1, 33.5) | 1130 | 14.6 (12.8, 16.5) | 469 | 27.8 (21.5, 34.0) | 692 | 12.8 (10.2, 15.4) |
| India 4 (Uddanam) | Kanchili | Rural | 148 | 11.3 (6.3, 16.2) | 169 | 15.8 (10.2, 21.4) | 71 | 13.0 (5.6, 20.5) | 83 | 13.9 (6.7, 21.1) |
| India 4 (Uddanam) | Kaviti | Rural | 102 | 19.7 (14.4, 24.9) | 110 | 20.0 (13.1, 26.9) | 52 | 12.5 (5.2, 19.7) | 65 | 14.0 (7.5, 20.5) |
| India 4 (Uddanam) | Mandasa | Rural | 95 | 24.2 (16.1, 32.3) | 105 | 18.5 (13.1, 24.0) | 57 | 18.3 (8.5, 28.0) | 54 | 12.5 (6.7, 18.3) |
| India 4 (Uddanam) | Palasa | Rural | 186 | 21.4 (15.5, 27.4) | 176 | 21.9 (15.5, 28.3) | 84 | 17.4 (9.2, 25.6) | 81 | 15.5 (8.5, 22.5) |
| India 4 (Uddanam) | Sompeta | Rural | 202 | 16.2 (11.1, 21.4) | 241 | 12.7 (8.4, 17.0) | 99 | 14.0 (6.9, 21.0) | 127 | 17.1 (10.3, 23.9) |
| India 4 (Uddanam) | V_kothuru | Rural | 250 | 14.7 (10.9, 18.5) | 281 | 13.7 (10.6, 16.8) | 101 | 11.4 (5.6, 17.3) | 121 | 11.6 (6.5, 16.7) |
| India 5 (Prakasam) | Kanigiri | Rural | 420 | 27.8 (23.7, 31.8) | 632 | 14.4 (11.8, 17.0) | 221 | 25.5 (20.0, 31.1) | 432 | 11.7 (8.3, 15.1) |
| Italy | Barga | Rural | 128 | 31.5 (23.9, 39.2) | 173 | 38.0 (31.2, 44.8) | 73 | 31.5 (21.8, 41.2) | 149 | 36.1 (29.3, 43.0) |
| Kenya | Muhoroni East | Urban | 138 | 8.1 (4.0, 12.3) | 122 | 20.5 (12.1, 28.9) | 113 | 8.6 (3.6, 13.6) | 104 | 21.6 (17.5, 25.6) |
| Kenya | Owaga | Rural | 113 | 10.3 (4.4, 16.1) | 129 | 16.1 (10.5, 21.7) | 88 | 8.5 (1.3, 15.8) | 98 | 17.9 (11.6, 24.2) |
| Kenya | Tonde | Rural | 114 | 3.6 (0.5, 6.7) | 119 | 7.2 (2.6, 11.9) | 94 | 3.5 (0.2, 6.7) | 100 | 8.2 (2.5, 13.8) |
| Malawi | Karonga | Rural | 271 | 17.6 (12.9, 22.3) | 375 | 14.5 (10.3, 18.8) | 214 | 15.0 (9.5, 20.4) | 309 | 13.8 (9.1, 18.5) |

| | | | | | | | | | | |
|---|---|---|---|---|---|---|---|---|---|---|
| Malawi | Lilongwe | Urban | 96 | 24.4 (13.9, 34.9) | 216 | 24.7 (18.1, 31.3) | 74 | 27.4 (16.2, 38.6) | 159 | 31.3 (18.0, 44.6) |
| Nepal | all | Rural | 1690 | 31.4 (29.2, 33.7) | 2790 | 44.1 (42.4, 45.9) | 1002 | 29.3 (26.6, 32.0) | 1981 | 43.2 (41.2, 45.2) |
| Nepal | all | Urban | 1602 | 40.0 (37.5, 42.6) | 2834 | 54.9 (53.0, 56.8) | 822 | 38.5 (35.3, 41.7) | 1777 | 53.8 (51.6, 56.1) |
| Nicaragua 1 | Chinandega (banana/sugarcane) | Rural | 155 | 32.9 (25.5, 40.3) | 176 | 6.5 (2.8, 10.3) | 104 | 25.1 (16.4, 33.8) | 111 | 2.2 (0, 4.6) |
| Nicaragua 1 | Chinandega (service) | Rural | 50 | 9.3 (1.3, 17.2) | 90 | 2.4 (0, 5.5) | 34 | 9.2 (0, 20.2) | 58 | 2.4 (0, 6.8) |
| Nicaragua 1 | Leon (coffee) | Rural | 40 | 8.6 (0, 17.8) | 37 | 5.6 (0, 14.4) | 30 | 11.2 (0, 22.8) | 26 | 0.0 (N/A) |
| Nicaragua 1 | Leon (fishing) | Rural | 76 | 31.2 (21.4, 40.9) | 90 | 11.6 (4.5, 18.8) | 55 | 24.5 (12.0, 36.9) | 73 | 6.4 (1.2, 11.7) |
| Nicaragua 1 | Leon (mining/subsistence farming) | Rural | 158 | 41.2 (34.2, 48.2) | 224 | 13.5 (8.8, 18.2) | 106 | 35.9 (26.1, 45.6) | 144 | 7.3 (1.7, 12.8) |
| Nicaragua 2 | Leon municipalities | Rural | 247 | 28.3 (23.0, 33.6) | 329 | 11.5 (8.0, 14.9) | 145 | 23.5 (16.8, 30.2) | 211 | 9.3 (4.3, 14.3) |
| Nicaragua 2 | Leon municipalities | Urban | 400 | 21.9 (18.4, 25.4) | 696 | 13.0 (10.8, 15.2) | 256 | 15.9 (11.2, 20.6) | 436 | 9.1 (6.5, 11.8) |
| Peru | Tumbes | Rural | 278 | 7.5 (5.0, 10.0) | 344 | 4.0 (2.0, 6.0) | 210 | 4.5 (2.1, 7.0) | 285 | 4.9 (2.0, 7.7) |
| Peru | Tumbes | Urban | 257 | 15.5 (11.7, 19.3) | 359 | 5.2 (3.2, 7.3) | 186 | 14.4 (9.8, 19.0) | 305 | 4.6 (2.3, 6.8) |
| Sri Lanka | Halambagaswewa | Rural | 242 | 33.6 (28.6, 38.6) | 497 | 29.1 (25.9, 32.4) | 136 | 31.7 (25.1, 38.2) | 336 | 26.5 (22.6, 30.4) |
| Sri Lanka | Lolugaswewa | Rural | 221 | 39.5 (33.2, 45.8) | 569 | 34.5 (31.4, 37.6) | 138 | 38.1 (31.3, 44.9) | 372 | 34.0 (30.3, 37.6) |
| Sri Lanka | Pothana | Rural | 194 | 26.5 (22.0, 31.0) | 497 | 30.3 (27.1, 33.6) | 115 | 26.3 (20.1, 32.5) | 330 | 29.6 (25.6, 33.6) |
| Sri Lanka | Puhudivula | Rural | 222 | 46.3 (40.6, 51.9) | 576 | 46.0 (42.5, 49.5) | 112 | 49.0 (42.7, 55.4) | 378 | 44.3 (39.9, 48.6) |
| Sri Lanka | Sangilikandarawa | Rural | 270 | 30.4 (25.5, 35.3) | 548 | 29.5 (26.2, 32.7) | 157 | 27.7 (21.6, 33.8) | 346 | 27.9 (23.8, 31.9) |
| Thailand | Bangkok | Urban | 381 | 18.8 (15.0, 22.6) | 1223 | 6.6 (5.4, 7.9) | 233 | 20.3 (15.4, 25.2) | 943 | 6.4 (5.0, 7.8) |
| Thailand | Central | Rural | 606 | 11.6 (9.4, 13.9) | 795 | 10.6 (8.6, 12.6) | 424 | 11.4 (8.7, 14.2) | 576 | 10.0 (7.7, 12.3) |
| Thailand | Central | Urban | 519 | 15.9 (12.9, 18.8) | 832 | 8.2 (6.6, 9.7) | 336 | 15.0 (11.5, 18.5) | 587 | 7.7 (6.0, 9.5) |
| Thailand | North | Rural | 668 | 11.0 (8.8, 13.2) | 730 | 7.4 (5.7, 9.2) | 407 | 10.1 (7.4, 12.7) | 498 | 7.2 (5.2, 9.2) |
| Thailand | North | Urban | 445 | 16.3 (12.5, 20.1) | 604 | 9.1 (7.0, 11.1) | 267 | 15.2 (10.8, 19.7) | 409 | 8.6 (6.2, 11.0) |
| Thailand | North East | Rural | 577 | 12.5 (10.5, 14.6) | 637 | 33.2 (29.0, 37.5) | 428 | 11.9 (9.6, 14.2) | 479 | 33.1 (28.5, 37.7) |
| Thailand | North East | Urban | 492 | 16.4 (14.0, 18.8) | 609 | 42.1 (37.2, 46.9) | 351 | 14.7 (12.0, 17.5) | 450 | 38.8 (33.7, 43.9) |
| Thailand | South | Rural | 549 | 9.9 (7.7, 12.0) | 678 | 4.3 (2.9, 5.6) | 366 | 10.0 (7.2, 12.7) | 493 | 4.0 (2.5, 5.6) |
| Thailand | South | Urban | 303 | 14.5 (11.1, 17.9) | 489 | 7.7 (5.6, 9.8) | 193 | 13.7 (9.5, 17.9) | 356 | 7.3 (4.8, 9.7) |
| USA | all | All | 1586 | 27.0 (24.9, 29.0) | 1787 | 17.9 (16.2, 19.5) | 925 | 28.2 (25.4, 30.9) | 1143 | 16.9 (14.8, 19.0) |

[a] age-standardised prevalence using WHO global population age weights; [b] only 41-60 years included; eGFR=creatinine-based estimated glomerular filtration rate; CI=confidence interval using normal approximation; N/A=no CI available due to zero estimate

Table S4: Lin's concordance correlation coefficient between eGFR measurements using creatinine and cystatin C, based on total sample

| Concordance between creatinine-based eGFR and: | England | India 3 (UDAY) | Kenya | Malawi | Peru |
|---|---|---|---|---|---|
| Cystatin C-based eGFR (CKD-EPI 2012) CCC (95% CI) | 0.53 (0.50-0.56) | 0.24 (0.21-0.26) | 0.44 (0.36-0.53) | 0.24 (0.18-0.29) | 0.49 (0.46-0.53) |
| Creatinine-cystatin-based eGFR (CKD-EPI 2012) CCC (95% CI) | N/A[a] | 0.55 (0.53-0.58) | 0.75 (0.69-0.81) | 0.82 (0.80-0.84) | 0.75 (0.73-0.77) |

eGFR=estimated glomerular filtration rate; CCC= Lin's concordance correlation coefficient; CI=confidence interval; [a] underlying cystatin C measurement not available

Table S5: Age-standardised[a] prevalence of creatinine- and cystatin C-based eGFR<60ml/min/1.73m$^2$ in complete sample (including those with diabetes, hypertension and heavy proteinuria), by sex, for ages 18-60 years with both creatinine and cystatin C measurements available

| Centre | Area | Rural / Urban | Men | | | | Women | | | |
|---|---|---|---|---|---|---|---|---|---|---|
| | | | n | Creatinine % (95% CI) | Cystatin C % (95% CI) | Creatinine-cystatin % (95% CI) | n | Creatinine % (95% CI) | Cystatin C % (95% CI) | Creatinine-cystatin % (95% CI) |
| England | all | Rural | 161 | 0.4 (0, 1.0) | 0.6 (0, 1.4) | -[b] | 223 | 2.2 (0.8, 3.7) | 0.4 (0, 0.9) | -[b] |
| England | all | Urban | 744 | 0.5 (0.1, 0.9) | 1.6 (0.7, 2.5) | -[b] | 1007 | 1.0 (0.6, 1.5) | 1.0 (0.5, 1.4) | -[b] |
| India 3 (UDAY)[c] | Sonipat | Rural | - | - | - | - | 253 | 0.0 (N/A) | 12.9 (7.3, 18.4) | 0.6 (0, 1.3) |
| India 3 (UDAY)[c] | Sonipat | Urban | - | - | - | - | 273 | 0.0 (N/A) | 14.1 (10.6, 17.6) | 2.8 (0.8, 4.8) |
| India 3 (UDAY)[c] | Vizag | Rural | - | - | - | - | 325 | 2.8 (1.2, 4.5) | 12.1 (8.7, 15.5) | 4.8 (2.7, 7.0) |
| India 3 (UDAY)[c] | Vizag | Urban | - | - | - | - | 244 | 0.0 (N/A) | 12.8 (8.1, 17.5) | 2.1 (0.1, 4.1) |
| Kenya | Muhoroni East | Urban | 4 | 0.0 (N/A) | 0.0 (N/A) | 0.0 (N/A) | 10 | 0.0 (N/A) | 0.0 (N/A) | 0.0 (N/A) |
| Kenya | Owaga | Rural | 44 | 0.0 (N/A) | 3.6 (0, 10.0) | 0.0 (N/A) | 50 | 0.0 (N/A) | 8.3 (3.2, 13.5) | 0.0 (N/A) |
| Kenya | Tonde | Rural | 26 | 0.0 (N/A) | 0.0 (N/A) | 0.0 (N/A) | 39 | 0.0 (N/A) | 10.6 (2.7, 18.5) | 0.0 (N/A) |
| Malawi | Karonga | Rural | 271 | 3.0 (0.7, 5.4) | 0.8 (0, 1.8) | 2.3 (0.3, 4.4) | 375 | 3.0 (1.0, 5.1) | 1.5 (0, 3.2) | 2.2 (0.3, 4.0) |
| Malawi | Lilongwe | Urban | 96 | 2.2 (0, 6.2) | 0.6 (0, 1.8) | 0.0 (N/A) | 216 | 4.9 (1.4, 8.4) | 0.0 (N/A) | 0.5 (0, 1.3) |
| Peru | Tumbes | Rural | 278 | 0.5 (0, 1.3) | 2.1 (0.7, 3.6) | 0.8 (0, 1.7) | 343 | 0.3 (0, 0.9) | 1.5 (0.2, 2.7) | 0.3 (0, 0.9) |
| Peru | Tumbes | Urban | 257 | 0.5 (0, 1.2) | 1.5 (0.2, 2.8) | 0.3 (0, 0.8) | 358 | 0.5 (0, 1.1) | 2.4 (1.0, 3.8) | 1.0 (0.0, 1.9) |

eGFR=estimated glomerular filtration rate; CI=Confidence interval using normal approximation; [a] age-standardised prevalence using WHO global population age weights; [b] not available as eGFR values supplied and exact age not available; [c] complete data not available as only participants without hypertension, diabetes and heavy proteinuria had cystatin C measured; N/A=no CI available due to zero estimate

Table S6: Sensitivity analysis for using age-adapted cut-offs for age-standardised[a] prevalence of eGFR<60ml/min/1.73m$^2$, compared to standard, in people without hypertension, diabetes or heavy proteinuria

| Centre | Area | Urbanicity | Men | | | Women | | |
|---|---|---|---|---|---|---|---|---|
| | | | n | Standard[b] | Age-adapted[c] | n | Standard[b] | Age-adapted[c] |
| England | all | Rural | 98 | 0.0 (N/A) | 2.8 (0, 6.5) | 169 | 1.8 (0.4, 3.2) | 5.0 (1.0, 9.1) |
| England | all | Urban | 515 | 0.1 (0, 0.4) | 2.9 (1.3, 4.4) | 759 | 0.8 (0.3, 1.4) | 2.1 (1.0, 3.2) |
| Guatemala | San Antonio Suchitepequez | Rural | 86 | 3.0 (0, 7.0) | 3.0 (0, 7.0) | 171 | 0.0 (N/A) | 0.4 (0, 1.2) |
| Guatemala | Tecpan | Rural | 83 | 0.0 (N/A) | 0.0 (N/A) | 152 | 0.0 (N/A) | 0.4 (0, 1.3) |
| India 1 (CARRS) | Chennai | Urban | 1161 | 0.5 (0.0, 0.9) | 0.5 (0.0, 0.9) | 1915 | 0.2 (0, 0.5) | 0.3 (0.0, 0.6) |
| India 1 (CARRS) | Delhi | Urban | 770 | 0.6 (0.1, 1.1) | 1.0 (0.3, 1.6) | 935 | 0.6 (0.0, 1.1) | 0.9 (0.2, 1.5) |
| India 2 (ICMR) | Delhi | Urban | 399 | 0.6 (0, 1.3) | 1.1 (0.2, 2.0) | 571 | 1.8 (0.6, 3.0) | 3.4 (1.8, 5.0) |
| India 2 (ICMR) | Faridabad | Rural | 380 | 1.9 (0.8, 3.1) | 3.0 (1.6, 4.3) | 520 | 1.5 (0.5, 2.5) | 5.5 (0, 11.3) |
| India 3 (UDAY) | Sonipat | Rural | 530 | 0.4 (0.0, 0.8) | 0.6 (0.1, 1.1) | 847 | 0.5 (0.1, 0.9) | 0.6 (0.2, 1.0) |
| India 3 (UDAY) | Sonipat | Urban | 586 | 0.6 (0.1, 1.1) | 1.1 (0.4, 1.8) | 768 | 0.3 (0, 0.6) | 0.3 (0, 0.7) |
| India 3 (UDAY) | Vizag | Rural | 696 | 6.5 (1.9, 11.0) | 6.9 (2.4, 11.4) | 933 | 2.6 (1.5, 3.7) | 4.0 (2.7, 5.3) |
| India 3 (UDAY) | Vizag | Urban | 469 | 0.4 (0, 1.1) | 1.6 (0.7, 2.6) | 692 | 0.5 (0, 1.1) | 0.6 (0, 1.3) |
| Italy | Barga | Rural | 73 | 1.2 (0, 3.5) | 1.2 (0, 3.5) | 149 | 0.0 (N/A) | 0.9 (0, 2.7) |
| Kenya | Muhoroni East | Urban | 113 | 0.0 (N/A) | 0.0 (N/A) | 104 | 0.0 (N/A) | 1.4 (0, 3.3) |
| Kenya | Owaga | Rural | 88 | 0.0 (N/A) | 0.8 (0, 2.4) | 98 | 2.4 (0, 5.8) | 2.4 (0, 5.8) |
| Kenya | Tonde | Rural | 94 | 0.0 (N/A) | 0.0 (N/A) | 100 | 0.0 (N/A) | 0.0 (N/A) |
| Malawi | Karonga | Rural | 214 | 1.4 (0, 3.5) | 2.9 (0.4, 5.5) | 309 | 2.8 (0.3, 5.3) | 3.1 (0.5, 5.6) |
| Malawi | Lilongwe | Urban | 74 | 3.1 (0, 8.6) | 4.5 (0, 10.4) | 159 | 4.0 (0.7, 7.2) | 6.8 (2.8, 10.8) |
| Nepal | all | Rural | 1002 | 0.5 (0.1, 1.0) | 2.4 (1.4, 3.5) | 1981 | 1.7 (1.1, 2.3) | 5.7 (4.7, 6.6) |
| Nepal | all | Urban | 822 | 0.3 (0.0, 0.7) | 2.0 (1.0, 3.0) | 1777 | 2.6 (1.9, 3.4) | 8.0 (6.7, 9.3) |
| Peru | Tumbes | Rural | 210 | 0.0 (N/A) | 0.0 (N/A) | 285 | 0.6 (0, 1.7) | 0.6 (0, 1.7) |
| Peru | Tumbes | Urban | 186 | 0.0 (N/A) | 0.0 (N/A) | 305 | 0.0 (N/A) | 0.0 (N/A) |
| Sri Lanka | Halambagaswewa | Rural | 136 | 3.9 (1.1, 6.8) | 12.6 (7.6, 17.6) | 336 | 1.3 (0.1, 2.6) | 9.6 (6.8, 12.4) |
| Sri Lanka | Lolugaswewa | Rural | 138 | 7.7 (3.9, 11.5) | 15.6 (10.8, 20.4) | 372 | 2.6 (1.0, 4.2) | 12.4 (9.4, 15.4) |
| Sri Lanka | Pothana | Rural | 115 | 0.7 (0, 1.9) | 5.6 (2.0, 9.2) | 330 | 0.7 (0, 1.6) | 8.1 (5.3, 10.9) |
| Sri Lanka | Puhudivula | Rural | 112 | 7.6 (3.8, 11.3) | 19.1 (13.6, 24.6) | 378 | 1.4 (0.2, 2.6) | 14.8 (11.3, 18.3) |
| Sri Lanka | Sangilikandarawa | Rural | 157 | 3.1 (0.6, 5.6) | 6.9 (3.4, 10.4) | 346 | 2.4 (0.8, 3.9) | 10.3 (7.3, 13.3) |
| USA | all | All | 925 | 0.6 (0.1, 1.0) | 2.5 (1.4, 3.5) | 1143 | 0.7 (0.2, 1.2) | 1.0 (0.4, 1.6) |

eGFR=estimated glomerular filtration rate; CI=Confidence interval using normal approximation; [a] age-standardised prevalence using WHO global population age weights; [b] standard cut-off of eGFR<60ml/min/1.73m$^2$; [c] age-adjusted cut-off of eGFR<60ml/min/1.73m$^2$ for age 40-60 and eGFR<75ml/min/1.73m$^2$ for age <4

Table S7: Sensitivity analysis for different eGFR equations. Age-standardised[a] prevalence of eGFR<60ml/min/1.73m$^2$ in people without hypertension, diabetes or heavy proteinuria, in population aged 18-60 years

| Centre | Area | Urbanicity | Men | | | | Women | | | |
|---|---|---|---|---|---|---|---|---|---|---|
| | | | n | CKD-EPI 2009[b] | CKD-EPI 2021 | CKD-MDRD[b] | n | CKD-EPI 2009[b] | CKD-EPI 2021 | CKD-MDRD[b] |
| Chile | Molina | Rural | 16 | - | 0.0 (N/A) | - | 16 | - | 0.0 (N/A) | - |
| Chile | Molina | Urban | 66 | - | 0.0 (N/A) | - | 66 | - | 0.5 (0, 1.6) | - |
| Ecuador | Miguelillo | Rural | 180 | 1.2 (0, 2.8) | - | - | 235 | 6.0 (2.2, 9.7) | - | - |
| England | all | Rural | 98 | 0.0 (N/A) | - | - | 169 | 1.8 (0.4, 3.2) | - | - |
| England | all | Urban | 515 | 0.1 (0, 0.4) | - | - | 759 | 0.8 (0.3, 1.4) | - | - |
| Guatemala | San Antonio Suchitepequez | Rural | 86 | 3.0 (0, 7.0) | 1.5 (0, 4.3) | 3.0 (0, 7.0) | 171 | 0.0 (N/A) | 0.0 (N/A) | 0.0 (N/A) |
| Guatemala | Tecpan | Rural | 83 | 0.0 (N/A) | 0.0 (N/A) | 0.0 (N/A) | 152 | 0.0 (N/A) | 0.0 (N/A) | 0.0 (N/A) |
| India 1 (CARRS) | Chennai | Urban | 1161 | 0.5 (0.0, 0.9) | 0.3 (0, 0.7) | 0.5 (0.0, 1.0) | 1915 | 0.2 (0, 0.5) | 0.1 (0, 0.2) | 0.5 (0.1, 0.9) |
| India 1 (CARRS) | Delhi | Urban | 770 | 0.6 (0.1, 1.1) | 0.4 (0, 0.8) | 1.3 (0.5, 2.1) | 935 | 0.6 (0.0, 1.1) | 0.4 (0.0, 0.7) | 1.1 (0.3, 1.9) |
| India 2 (ICMR) | Delhi | Urban | 399 | 0.6 (0, 1.3) | 0.1 (0, 0.4) | 2.2 (0.8, 3.6) | 571 | 1.8 (0.6, 3.0) | 1.1 (0.2, 2.0) | 3.6 (1.9, 5.2) |
| India 2 (ICMR) | Faridabad | Rural | 380 | 1.9 (0.8, 3.1) | 1.6 (0.6, 2.7) | 2.9 (1.6, 4.3) | 520 | 1.5 (0.5, 2.5) | 0.9 (0.2, 1.7) | 6.3 (0.4, 12.2) |
| India 3 (UDAY) | Sonipat | Rural | 530 | 0.4 (0.0, 0.8) | 0.2 (0, 0.6) | 0.6 (0.1, 1.1) | 847 | 0.5 (0.1, 0.9) | 0.5 (0.1, 0.9) | 1.1 (0.5, 1.7) |
| India 3 (UDAY) | Sonipat | Urban | 586 | 0.6 (0.1, 1.1) | 0.5 (0.0, 0.9) | 0.9 (0.3, 1.6) | 768 | 0.3 (0, 0.6) | 0.1 (0, 0.4) | 1.0 (0.4, 1.7) |
| India 3 (UDAY) | Vizag | Rural | 696 | 6.5 (1.9, 11.0) | 5.9 (1.4, 10.4) | 7.7 (3.2, 12.3) | 933 | 2.6 (1.5, 3.7) | 2.3 (1.3, 3.3) | 4.7 (3.4, 6.0) |
| India 3 (UDAY) | Vizag | Urban | 469 | 0.4 (0, 1.1) | 0.4 (0, 1.1) | 0.9 (0.1, 1.8) | 692 | 0.5 (0, 1.1) | 0.5 (0, 1.1) | 1.5 (0.5, 2.6) |
| India 4 (Uddanam) | Kanchili | Rural | 71 | 6.9 (1.0, 12.9) | - | - | 83 | 1.1 (0, 3.2) | - | - |
| India 4 (Uddanam) | Kaviti | Rural | 52 | 6.7 (1.3, 12.2) | - | - | 65 | 8.0 (2.0, 14.1) | - | - |
| India 4 (Uddanam) | Mandasa | Rural | 57 | 13.7 (4.8, 22.6) | - | - | 54 | 5.6 (0.4, 10.7) | - | - |
| India 4 (Uddanam) | Palasa | Rural | 84 | 6.8 (0.1, 13.4) | - | - | 81 | 4.6 (0.5, 8.7) | - | - |
| India 4 (Uddanam) | Sompeta | Rural | 99 | 8.2 (2.2, 14.3) | - | - | 127 | 5.8 (1.1, 10.6) | - | - |
| India 4 (Uddanam) | V_kothuru | Rural | 101 | 2.1 (0, 4.8) | - | - | 121 | 7.5 (3.2, 11.9) | - | - |
| India 5 (Prakasam) | Kanigiri | Rural | 221 | - | 2.5 (0.3, 4.6) | - | 221 | - | 1.9 (0.7, 3.1) | - |
| Italy | Barga | Rural | 73 | 1.2 (0, 3.5) | 1.2 (0, 3.5) | 3.1 (0, 6.5) | 149 | 0.0 (N/A) | 0.0 (N/A) | 1.7 (0.1, 3.3) |
| Kenya | Muhoroni East | Urban | 113 | 0.0 (N/A) | 0.0 (N/A) | 0.0 (N/A) | 113 | 0.0 (N/A) | 0.0 (N/A) | 17.4 (15.9, 18.9) |
| Kenya | Owaga | Rural | 88 | 0.0 (N/A) | 0.0 (N/A) | 0.0 (N/A) | 98 | 2.4 (0, 5.8) | 2.4 (0, 5.8) | 2.4 (0, 5.8) |
| Kenya | Tonde | Rural | 94 | 0.0 (N/A) | 0.0 (N/A) | 0.0 (N/A) | 100 | 0.0 (N/A) | 0.0 (N/A) | 0.0 (N/A) |
| Malawi | Karonga | Rural | 214 | 1.4 (0, 3.5) | 1.4 (0, 3.5) | 2.5 (0, 5.3) | 309 | 2.8 (0.3, 5.3) | 2.8 (0.3, 5.3) | 3.7 (1.0, 6.5) |
| Malawi | Lilongwe | Urban | 74 | 3.1 (0, 8.6) | 3.1 (0, 8.6) | 4.5 (0, 10.4) | 159 | 4.0 (0.7, 7.2) | 4.0 (0.7, 7.2) | 5.7 (1.9, 9.5) |
| Nepal | all | Rural | 1002 | - | - | 0.5 (0.1, 1.0) | 1981 | - | - | 1.7 (1.1, 2.3) |
| Nepal | all | Urban | 822 | - | - | 0.3 (0.0, 0.7) | 1777 | - | - | 2.6 (1.9, 3.4) |
| Nicaragua 1 | Chinandega (banana/sugarcane) | Rural | 104 | - | - | 13.6 (6.3, 20.9) | 111 | - | - | 0.0 (N/A) |

| | | | | | | | | | |
|---|---|---|---|---|---|---|---|---|---|
| Nicaragua 1 | Chinandega (service) | Rural | 34 | - | - | 0.0 (N/A) | 58 | - | - | 0.0 (N/A) |
| Nicaragua 1 | Leon (coffee) | Rural | 30 | - | - | 8.6 (0, 19.1) | 26 | - | - | 0.0 (N/A) |
| Nicaragua 1 | Leon (fishing) | Rural | 55 | - | - | 6.4 (0, 13.0) | 73 | - | - | 0.0 (N/A) |
| Nicaragua 1 | Leon (mining/ subsistence farming) | Rural | 106 | - | - | 12.1 (4.5, 19.6) | 144 | - | - | 2.6 (0, 6.6) |
| Nicaragua 2 | Leon municipalities | Rural | 145 | 9.4 (4.4, 14.3) | - | - | 211 | 2.5 (0, 5.4) | - | - |
| Nicaragua 2 | Leon municipalities | Urban | 256 | 6.6 (3.0, 10.1) | - | - | 436 | 2.4 (1.0, 3.9) | - | - |
| Peru | Tumbes | Rural | 210 | 0.0 (N/A) | 0.0 (N/A) | 0.4 (0, 1.2) | 285 | 0.6 (0, 1.7) | 0.0 (N/A) | 1.2 (0, 2.8) |
| Peru | Tumbes | Urban | 186 | 0.0 (N/A) | 0.0 (N/A) | 0.4 (0, 1.2) | 305 | 0.0 (N/A) | 0.0 (N/A) | 0.6 (0, 1.5) |
| Sri Lanka | Halambagaswewa | Rural | 136 | 3.9 (1.1, 6.8) | - | - | 336 | 1.3 (0.1, 2.6) | - | - |
| Sri Lanka | Lolugaswewa | Rural | 138 | 7.7 (3.9, 11.5) | - | - | 372 | 2.6 (1.0, 4.2) | - | - |
| Sri Lanka | Pothana | Rural | 115 | 0.7 (0, 1.9) | - | - | 330 | 0.7 (0, 1.6) | - | - |
| Sri Lanka | Puhudivula | Rural | 112 | 7.6 (3.8, 11.3) | - | - | 378 | 1.4 (0.2, 2.6) | - | - |
| Sri Lanka | Sangilikandarawa | Rural | 157 | 3.1 (0.6, 5.6) | - | - | 346 | 2.4 (0.8, 3.9) | - | - |
| Thailand | Bangkok | Urban | 233 | 1.2 (0, 2.7) | - | - | 943 | 0.5 (0, 1.1) | - | - |
| Thailand | Central Rural | Rural | 424 | 0.2 (0, 0.4) | - | - | 576 | 1.0 (0.1, 1.8) | - | - |
| Thailand | Central Urban | Urban | 336 | 0.0 (N/A) | - | - | 587 | 0.2 (0, 0.5) | - | - |
| Thailand | North Rural | Rural | 407 | 0.4 (0, 0.9) | - | - | 498 | 0.5 (0, 1.3) | - | - |
| Thailand | North Urban | Urban | 267 | 0.7 (0, 1.9) | - | - | 409 | 0.3 (0, 0.7) | - | - |
| Thailand | Northeast Rural | Rural | 428 | 0.6 (0.1, 1.2) | - | - | 479 | 0.3 (0, 0.7) | - | - |
| Thailand | Northeast Urban | Urban | 351 | 0.2 (0, 0.5) | - | - | 450 | 0.1 (0, 0.4) | - | - |
| Thailand | South Rural | Rural | 366 | 0.2 (0, 0.6) | - | - | 493 | 0.2 (0, 0.5) | - | - |
| Thailand | South Urban | Urban | 193 | 1.1 (0, 2.4) | - | - | 356 | 0.7 (0, 1.4) | - | - |
| USA | all | All | 925 | 0.6 (0.1, 1.0) | 0.3 (0.0, 0.7) | 1.7 (0.9, 2.6) | 1143 | 0.7 (0.2, 1.2) | 0.6 (0.1, 1.0) | 1.8 (1.0, 2.5) |

eGFR=estimated glomerular filtration rate; CI=Confidence interval using normal approximation; [a] age-standardised prevalence using WHO global population age weights; [b] without race adjustment

Table S8: Linear regression associations between prevalence of eGFR<60ml/min/1.73m$^2$ in people without hypertension, diabetes or heavy proteinuria, and sampling characteristics

| Characteristic | Median (range) of values | Associations[a] | | |
|---|---|---|---|---|
| | | All centres (n=108[d]) estimate (*P*-value) | Affected centres[b] (n=36) estimate (*P*-value) | Unaffected centres[c] (n=72[d]) estimate (*P*-value) |
| Response rate (per 1% absolute increase) | 85% (37% - 98%) | 0.01% (0.67) | 0.01% (0.72) | -0.003% (0.59) |
| Study year (per year increase) | 2017 (2007 - 2023) | -0.06% (0.39) | -0.18% (0.11) | 0.002% (0.91) |
| Proportion of males (per 1% absolute increase) | 43% (24% - 53%) | 0.04% (0.41) | 0.15% (0.04) | -0.02 (0.12) |

eGFR=estimated glomerular filtration rate; [a] effect on absolute percentage of prevalence after adjusting for sex and urban/rural; [b] affected centres have moderate-high prevalence of >=2%; [c] unaffected centres have low prevalence of <2%; [d] there are four fewer in the response rate sample due to missing response rates
